# Serotype-Specific Epidemiological Patterns of Inapparent versus Symptomatic Primary Dengue Virus Infections: A 17-year cohort study in Nicaragua

**DOI:** 10.1101/2024.04.05.24305281

**Authors:** Sandra Bos, Jose Victor Zambrana, Elias Duarte, Aaron L. Graber, Julia Huffaker, Carlos Montenegro, Lakshmanane Premkumar, Aubree Gordon, Guillermina Kuan, Angel Balmaseda, Eva Harris

**Author notes:** These authors contributed equally to this work (Sandra Bos, Jose Victor Zambrana).

## Abstract

**Background:** Dengue is the most prevalent mosquito-borne viral disease and a major public health problem worldwide. Most primary infections with the four dengue virus serotypes (DENV1-4) are inapparent; nonetheless, whether the distribution of symptomatic versus inapparent infections by serotype varies remains unknown. Here, we present (1) the evaluation of a multiplex DENV1-4 envelope domain III multiplex microsphere-based assay (EDIII-MMBA) to serotype inapparent primary infections and (2) its application leveraging 17 years of prospective sample collection from the Nicaraguan Pediatric Dengue Cohort Study (PDCS).

**Methods:** First, we evaluated the performance of the EDIII-MMBA with samples characterized by RT-PCR or focus reduction neutralization test. Next, we analyzed 46% (N=574) of total inapparent primary DENV infections in the PDCS with the EDIII-MMBA to evaluate the epidemiology of inapparent infections. Remaining infections were inferred using stochastic imputation, taking year and neighborhood into account. Infection incidence and percentage of inapparent, symptomatic, and severe infections were analyzed by serotype.

**Findings:** The EDIII-MMBA demonstrated excellent overall accuracy (100%, 95·8-100%) for serotyping symptomatic and inapparent primary DENV infections when evaluated against gold-standard serotyping methods. We found that a significant majority of primary infections were inapparent, with DENV3 exhibiting the highest likelihood of symptomatic and severe primary infections (Pooled OR compared to DENV1 = 2·13, 95% CI 1·28-3·56, and 6·75, 2·01-22·62, respectively), whereas DENV2 was similar to DENV1 in both analyses. Significant within- and between-year variation in serotype distribution between symptomatic and inapparent infections and circulation of serotypes undetected in symptomatic cases were observed in multiple years.

**Interpretation:** Our study indicates that case surveillance skews the perceived epidemiological footprint of DENV. We reveal a more complex and intricate pattern of serotype distribution in inapparent infections. The significant differences in infection outcomes by serotype emphasizes the need for vaccines with balanced immunogenicity and efficacy across serotypes.

**Funding:** NIH/NIAID P01AI106695, U01AI153416

**Research in context:** *Evidence before this study:* We conducted a search in PubMed for studies published up to February 2024. Keywords included “dengue virus” and “DENV” in combination with “inapparent infections”, “asymptomatic infections”, “primary infections by serotype”, “FoI by serotype”, “force of infection”, “force of infection by serotype”, and identified a significant gap in the current understanding of dengue epidemiology. Despite acknowledging the high prevalence of inapparent DENV infections in endemic regions, previous research has focused primarily on symptomatic infections, potentially biasing our understanding of the DENV epidemiological landscape and hindering our capacity to determine the complete disease spectrum of the different DENV serotypes. While cross-sectional studies have provided preliminary insights into this gap, there is a need for more comprehensive and detailed serotype-specific insights to evaluate the epidemiological impact of inapparent infections. The lack of comprehensive characterization of inapparent infections reflects methodological challenges, particularly the need for prospective cohort studies designed to capture and accurately serotype these infections. Moreover, the reliance on labor-intensive and low-throughput antibody neutralization assays for serotyping, despite their accuracy, has constrained high-throughput analysis required for large-scale epidemiological studies.

*Added value of this study:* Our study, spanning 17 years of prospective cohort data in Nicaragua, addresses this bottleneck in dengue research by providing a detailed examination of primary inapparent infections. The introduction of a novel envelope domain III (EDIII) multiplex microsphere-based assay for DENV serotyping represents a significant methodological advance, offering an efficient, scalable alternative for large epidemiological studies. A key contribution of our study is the intricate pattern of serotype distribution among inapparent infections. In contrast to the serotype predominance observed in symptomatic infections, inapparent infections exhibit a complex landscape with co-circulation of multiple DENV serotypes, including serotypes undetected in symptomatic surveillance in multiple years. Our systematic documentation of the entire disease spectrum provides unprecedented insights into the serotype-specific disease burden in primary infection, including the proportion of symptomatic versus inapparent infection and its temporal variations, thus providing a more complete picture of DENV epidemiology than has been available to date. Notably, we demonstrate striking differences in disease severity by serotype, with DENV3 infections being significantly more symptomatic and more severe compared to DENV1 and DENV2, the latter displaying the highest rate of inapparent infection.

*Implications of all the available evidence:* Our research challenges prior assumptions by demonstrating that inapparent and symptomatic primary DENV infections present distinct epidemiological profiles, revealing that the epidemiological footprint of DENV is broader and more nuanced than previously recognized through symptomatic cases alone. These findings underscore the utility for continuous and comprehensive surveillance systems that capture both symptomatic and inapparent infections to accurately assess the epidemiological burden of DENV and inform public health interventions. Additionally, they provide critical insight for enhancing the accuracy of predictive DENV transmission modeling. Furthermore, the marked differences in infection outcomes by serotype emphasize the need for serotype-informed public health strategies. This nuanced understanding is pivotal for the crafting of targeted interventions, vaccine development and vaccination strategies, and efficient resource allocation, ultimately contributing to the global effort to mitigate the impact of dengue.

## Introduction

Dengue, caused by the four dengue virus serotypes (DENV1-4), is the most prevalent mosquito-borne viral disease in humans and a significant global health threat.^1^ Infection outcomes range from inapparent/subclinical infection to severe, potentially fatal disease. The global dissemination of DENV, alongside the co-circulation of its serotypes, has increased both the incidence and severity of the disease in recent decades.^1, 2^ This epidemiological shift emphasizes the need for a comprehensive understanding of the circulation and interactions among the different serotypes and their implications for disease outcomes.

Infection with one of the four antigenically related DENV serotypes leads to the production of type-specific antibodies as well as cross-reactive antibodies that can bind to other serotypes. These antibodies can either protect or increase the risk of subsequent symptomatic or severe DENV infection in both natural infections and vaccines.^3–5^ In this context, one factor that can influence clinical outcome is the infecting serotype.^5, 6^ Further, an association between the order of DENV serotype infection and clinical outcome has been reported,^6^ suggesting that the initial infecting serotype may modulate infection outcomes.^5, 7, 8^

While the majority of primary DENV infections are inapparent, they significantly contribute to the force of infection, transmission, and overall epidemiological burden.^9, 10^ However, accurately assessing this burden, understanding the complete disease spectrum associated with each serotype, and predicting the risk of severe disease outcomes upon subsequent infection have been challenging due to the lack of characterization of inapparent infections. This knowledge gap stems from methodological limitations, particularly the need for prospective cohort studies designed to capture and serotype inapparent infection, compounded by the low-throughput nature of neutralization assays that are used to identify serotype in inapparent infections. Consequently, prior research has predominantly focused on symptomatic infections, potentially skewing our understanding of the DENV epidemiological landscape.

To address this bottleneck in dengue research, we leveraged 17 years of prospective sample collection from participants enrolled in our pediatric cohort in Nicaragua and demonstrated the hidden contribution of inapparent infection to both the overall and serotype-specific DENV burden in Nicaragua between 2004 and 2021. To achieve this, we used an envelope domain III multiplex microsphere-based assay (EDIII-MMBA), previously validated to differentiate DENV and Zika virus (ZIKV) infections and determine the number of prior DENV infections, to serotype inapparent DENV infection.^11^ We then examined the distribution of DENV serotypes in inapparent versus symptomatic primary infections, the proportion of inapparent infections per serotype, and their temporal dynamics in the PDCS, together revealing the full disease spectrum associated with primary DENV infection by serotype.

## Methods

### Study design and participants

In this study, we first evaluated the performance of the EDIII-MMBA to serotype primary DENV infections and subsequently used it to characterize the serotype of inapparent primary infections and evaluate the epidemiological and clinical landscape of primary DENV infections in Nicaragua. To do so, we leveraged serum samples collected from 2004 to 2021 from our ongoing Pediatric Dengue Cohort Study (PDCS),^12^ which has followed ∼4,000 active participants aged 2-17 years old since 2004. The PDCS is conducted at the Sócrates Flores Vivas Health Center located in District 2 of Managua, Nicaragua. Dengue cases are detected by enhanced passive surveillance, and healthy samples are collected annually in March-April, which allows the detection of inapparent infections. Ethics statements, description of the PDCS and participants, case definitions, and severity classifications are described in the Supplementary Appendix.^12^ In this study, we included DENV-naïve participants at enrollment (N=5931) and followed them until they experienced a primary DENV infection (N=1626). Participants with DENV or ZIKV immunity at enrollment were excluded (N=5824).

### Procedures

A primary DENV infection was defined among individuals who entered the cohort DENV-naïve and ZIKV-naïve as measured by DENV iELISA and ZIKV NS1 blockade-of-binding ELISA and who maintained this status until their seroconversion to DENV. Symptomatic DENV infections were confirmed by detection of DENV RNA by RT-PCR and/or virus isolation in the acute phase sample and/or seroconversion detected in paired acute- and convalescent-phase samples by DENV IgM capture ELISA or iELISA. Inapparent primary DENV infections were detected by seroconversion between 2 consecutive annual samples in the absence of any documented dengue case, according to the case definition, in the intervening year. Disease severity of symptomatic DENV infections was classified according to the 2009 WHO Dengue Guidelines.^15^ Procedures, case definitions, and assays are explained in detail in the Supplementary Appendix.

The “gold standard” to serotype inapparent infections is the DENV1-4 neutralization assay, which is both time- and sample-intensive. To address these limitations, we evaluated the performance of a multiplex assay containing EDIII of DENV1-4 and ZIKV conjugated to avidin-coated microspheres via an Avi-Tag,^11^ the EDIII-MMBA, to serotype primary infections. The EDIII-MMBA was evaluated against RT-PCR and focus reduction neutralization test (FRNT) using 156 late convalescent samples from the annual sample collection (3-9 months post-infection) that constituted the evaluation set (86 inapparent infections and 70 symptomatic infections selected by convenience sampling). See Supplementary Appendix for details. Of the 1244 inapparent primary infections that occurred from 2004 to 2021 as captured by the DENV iELISA, 574 (46%) were processed by the EDIII-MMBA assay to study the epidemiology of primary DENV infection in the PDCS. Selection of this subset for screening by the EDIII-MMBA was performed by random selection of 40% of primary DENV-infected individuals each year, and when there were less than 20 inapparent primary infections, 100% were selected, restricted to those with serum/plasma available from the following annual sample collection (3-9 months post-infection). This sample selection yielded samples from 478 individuals (87%); to improve precision, we included 76 inapparent samples (13%; excluding 10 ZIKV infections) from the evaluation subset.

### Outcomes and endpoints

Outcomes included inapparent, symptomatic, severe, and overall primary DENV infections as defined in the Procedures section and Supplementary Appendix. Our primary endpoints included the ratio of symptomatic versus inapparent and severe versus non-severe DENV infections by serotype. Our secondary analyses include a year-by-year analysis of the relative distribution of DENV serotypes in inapparent versus symptomatic DENV infections, the percent of symptomatic DENV infections by serotype over time by serotype, and the overall infection incidence by serotype over time.

### Statistical methods

The accuracy of the EDIII-MMBA assay to serotype primary DENV infections was evaluated by comparing it to gold standards: RT-PCR (symptomatic) and FRNT (inapparent). Sensitivity, specificity, and accuracy, calculated as the sum of the true positives and true negatives divided by the total number of samples evaluated, were calculated for each serotype (DENV1-4) and Zika virus (ZIKV) across all test combinations. We calculated 95% confidence intervals (95% CI) using the exact binomial method for all the performance measures.

To determine population-level infection parameters over time by DENV serotype, including percent infected among the DENV-naïve population and percent of severe, symptomatic and inapparent primary infections, we addressed missingness of serotype in both inapparent and symptomatic primary DENV infections by employing multiple imputation with stochastic Multivariate Imputation by Chained Equations (MICE). A polytomous logistic regression model, incorporating epidemic year and neighborhood as predictors, was used to impute missing serotypes. We ran independent imputation models for inapparent and symptomatic infections, running 1000 simulations with 10 iterations for both models. The resulting imputed datasets were merged for analysis, pooling all results across simulations. To understand the temporal trends of infected individuals by outcome (symptomatic vs. inapparent) for each serotype, we plotted the pooled counts and percentages across imputations over time. Additionally, we calculated the relative distribution of serotypes by outcomes over time.

For our primary endpoints, we analyzed the ratio of symptomatic and severe infections out of the total infections specific to each serotype using separate logistic regression models by symptomatic and severe infections, adjusting by serotype and controlling by epidemic year. As secondary endpoints, we analyzed the within-year relative distribution of serotypes by inapparent and symptomatic infections by employing Fisher’s exact test to both serotyped and imputed data. Additionally, we analyzed changes in the percent symptomatic infections by DENV serotype over time by applying logistic regression models measuring the likelihood of symptomatic infection by epidemic year and stratifying by serotype. Finally, we analyzed the infection incidence proportion by serotype by applying intercept-only logistic regression models measuring the likelihood of infection stratifying by each serotype. A conceptual framework of our assumptions about the relationship between the variables used in our analyses is shown in the Supplemental Appendix. A validation of our final imputation model and various sensitivity analyses on our primary endpoints were performed (Supplementary Appendix). To account for the imputed data, we pooled the analyses across simulations using Rubin’s rule to obtain estimates and 95% CI for the model coefficients. All statistical analyses and data imputation were executed in R (version 4.3.2), utilizing the ‘mice’ package (version 3.16.0).

## Results

In this study, we analyzed primary DENV infection data from the PDCS from 2004 to 2021, involving a median of 1536 (IQR = 1425-1861) DENV-naïve participants followed annually (Table S1). Notably, this period includes the ZIKV epidemic in 2016, which significantly dampened DENV circulation in Nicaragua in 2016-2018. We first evaluated the count and incidence proportion of primary infections (figure 1A-B). Over the 17-year period, we observed significant year-to-year fluctuations in the number of primary infections, ranging from 6 in 2018 to 289 in 2019, with a median annual incidence proportion of 4% (IQR = 1-9%) within the study population (Table S1). Prior to the emergence of ZIKV, a median of 105 (IQR = 72-124) primary infections was recorded annually, which dropped to 9 (IQR = 7-12) from 2016 to 2018, before a significant resurgence of DENV primary infections in 2019 (N=287). Out of the 1626 total primary infections we studied, 382 were symptomatic, while 1244 were inapparent. We found that a median of 77% (IQR = 68-86) of primary infections were inapparent, although this percentage varied significantly across epidemic years (figure 1C).

**Figure 1.**
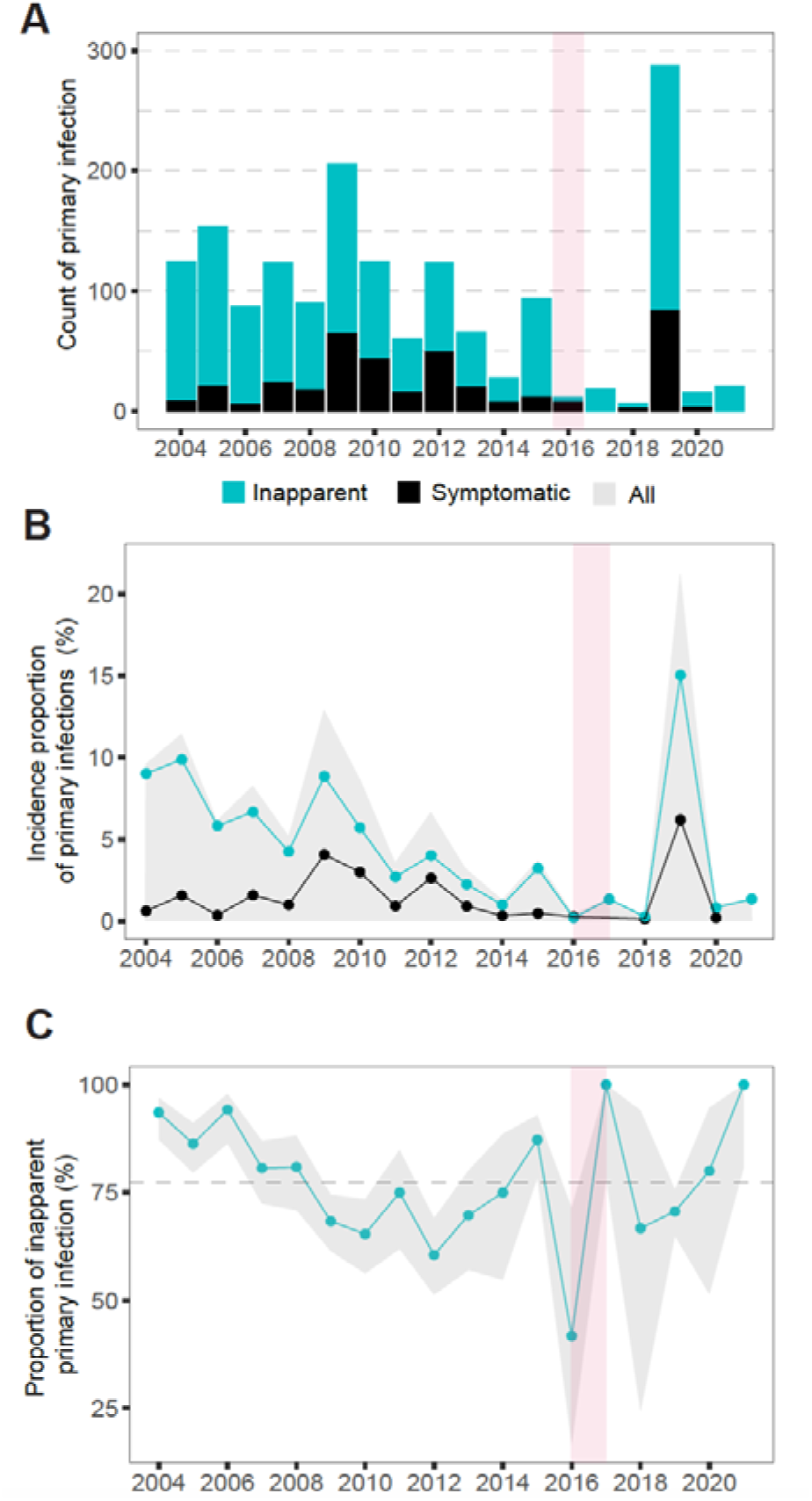
Overall symptomatic and inapparent DENV infections in the PDCS study from 2004 to 2021. **(A)** Yearly counts of DENV primary infections by symptomatic or inapparent status, as indicated. **(B)** Yearly cumulative incidence proportion of primary inapparent and symptomatic DENV infections. **(C)** Yearly percent of inapparent primary DENV infections. The pink shaded area in 2016 is the period when ZIKV circulated in the PDCS. The gray area in panel B covers the pooled incidence proportions of overall primary DENV infections by serotype. Dashed line in panel C is the yearly average of inapparent primary DENV infections across years, and gray area represents the confidence interval.

Considering the predominance of inapparent primary infections, serotyping these infections is critical to evaluate the epidemiological burden and virulence associated with each serotype. Thus, we evaluated the performance of the EDIII-MMBA to serotype primary infections. Exceptional performance (100% sensitivity, specificity, and accuracy) was observed when evaluated against both RT-PCR and FRNT for DENV1-4 and ZIKV (Table 1). Concordance of EDIII-MMBA values with RT-PCR and FRNT is shown in figure 2.

**Figure 2.**
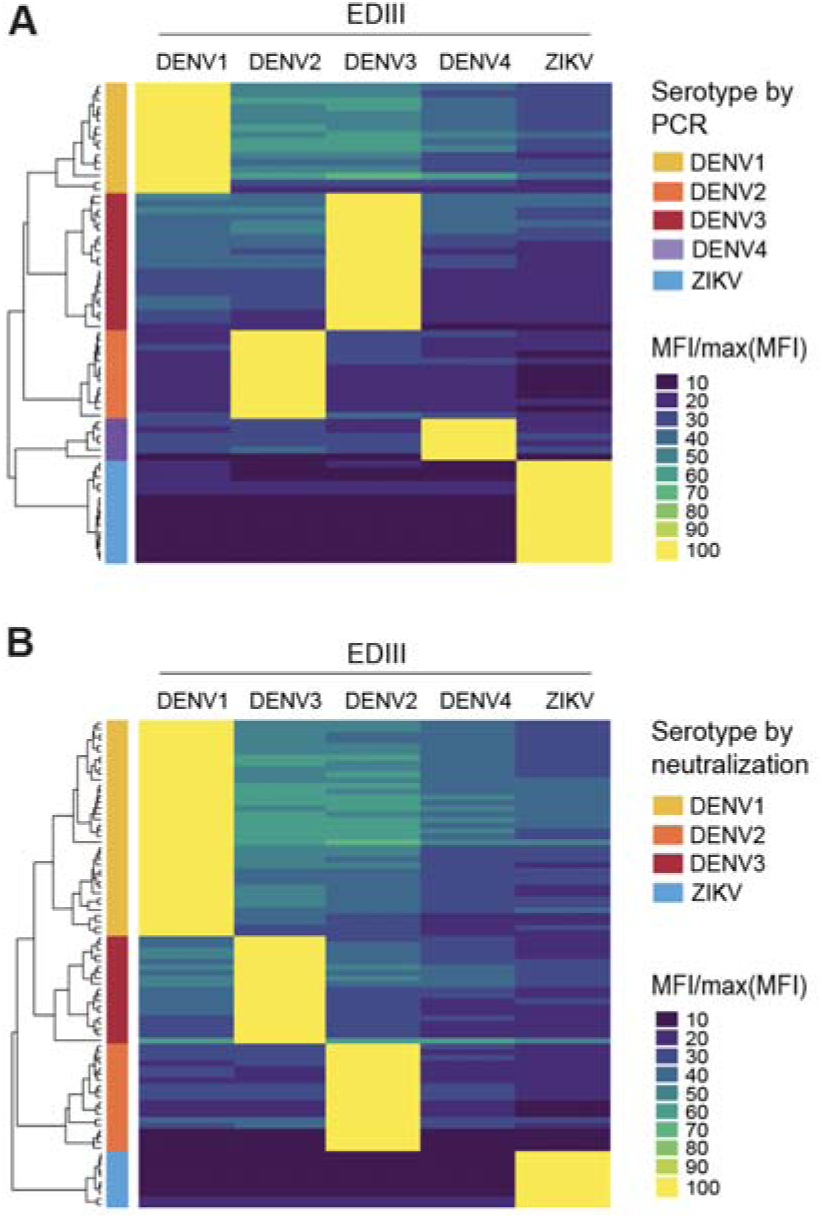
Concordance of relative EDIII-MMBA values from participants with primary DENV infection with values from RT-PCR and neutralization assay. **(A)** Symptomatic primary DENV infections confirmed by RT-PCR, and **(B)** inapparent primary DENV infections were confirmed by FRNT. Within the heatmaps, each cell indicates the Mean Fluorescence Intensity (MFI). MFI values per participant were transformed by dividing the MFI of each EDIII antigen by the highest MFI observed in the set. Adjacent to each heatmap, dendrograms display the hierarchical clustering of individuals based on Euclidean distances calculated for the normalized MFI values. On the right of the dendrograms, a color-coded column classifies individuals according to the “gold standard” assay results, providing a reference for true infection status.

**Table 1.** EDIII-MMBA performance.

|  | DENV1 | DENV2 | DENV3 | DENV4 | ZIKV |
| --- | --- | --- | --- | --- | --- |
| **RT-PCR vs EDIII-MMBA** |  |  |  |  |  |
| N | 70 | 70 | 70 | 70 | 70 |
| N by characteristic | TP<sup>1</sup> = 16 | TP = 13 | TP = 20 | TP = 6 | TP = 15 |
|  | FP<sup>2</sup> = 0 | FP = 0 | FP = 0 | FP = 0 | FP = 0 |
|  | FN<sup>3</sup> = 0 | FN = 0 | FN = 0 | FN = 0 | FN = 0 |
|  | TN<sup>4</sup> = 54 | TN = 57 | TN = 50 | TN = 64 | TN = 55 |
| Gold standard positive | 16 | 13 | 20 | 6 | 15 |
| Gold standard negative | 54 | 57 | 50 | 64 | 55 |
| Sensitivity | 100<br>(79.4 - 100) | 100<br>(75.3 - 100) | 100<br>(83.2 - 100) | 100<br>(54.1 - 100) | 100<br>(78.2 - 100) |
| Specificity | 100<br>(93.4 - 100) | 100<br>(93.7 - 100) | 100<br>(92.9 - 100) | 100<br>(94.4 - 100) | 100<br>(93.5 - 100) |
| Diagnostic accuracy | 100<br>(94.9 - 100) | 100<br>(94.9 - 100) | 100<br>(94.9 - 100) | 100<br>(94.9 - 100) | 100<br>(94.9 - 100) |
| **FRNT vs EDIII-MMBA** |  |  |  |  |  |
| N | 86 | 86 | 86 | - | 86 |
| N by characteristic | TP<sup>1</sup> = 38 | TP = 19 | TP = 19 | - | TP = 10 |
|  | FP<sup>2</sup> = 0 | FP = 0 | FP = 0 | - | FP = 0 |
|  | FN<sup>3</sup> = 0 | FN = 0 | FN = 0 | - | FN = 0 |
|  | TN<sup>4</sup> = 48 | TN = 67 | TN = 67 | - | TN = 76 |
| Gold standard positive | 38 | 19 | 19 | - | 10 |
| Gold standard negative | 48 | 67 | 67 | - | 76 |
| Sensitivity | 100<br>(90.7 - 100) | 100<br>(82.4 - 100) | 100<br>(82.4 - 100) | - | 100<br>(69.2 - 100) |
| Specificity | 100<br>(92.6 - 100) | 100<br>(94.6 - 100) | 100<br>(94.6 - 100) | - | 100<br>(95.3 - 100) |
| Diagnostic accuracy | 100<br>(95.8 - 100) | 100<br>(95.8 - 100) | 100<br>(95.8 - 100) | - | 100<br>(95.8 - 100) |

<sup>1</sup>TP: True positive ; <sup>2</sup>FP: False positive ; <sup>3</sup>FN: False Negative ; <sup>4</sup>TN: True Negative.

The EDIII-MMBA was conducted on 574 out of 1244 (46%) primary inapparent infections recorded in the PDCS between 2004 and 2021, of which 425 (74%) were successfully serotyped. Twenty-five infections (4%) were excluded due to either invalid serotyping (N=21) (see Supplementary Appendix) or ZIKV infection (N=14). Anti-EDIII antibody levels below the detection limit (1:50) were observed in 114 (19%) samples. This discrepancy might be due to the difference in the type of antibody measured (anti-EDIII IgG), compared to the iELISA used initially to detect infection (total Ig primarily directed towards anti-pr and anti-E fusion loop antibodies).^3^ Using the total number of samples screened, we performed an analysis of the capacity of the EDIII-MMBA to provide valid serotyping of primary infection samples as detected by iELISA (n=564). Including the 114 samples with no detectable anti-EDIII antibodies and the 21 samples with invalid serotyping, the sensitivity of the EDIII-MMBA was 76·5%, compared to 95·4% when only participants with anti-EDIII antibodies were included (n=460). The breakdown of serotyped and non-serotyped infections, along with a flowchart illustrating the screening and serotyping process, are provided in Table S2 and figure S1. Missing serotype data on the remaining inapparent and symptomatic infections were imputed using the results of EDIII-MMBA and RT-PCR, respectively (figure S2). The convergence of our final imputation models was assessed using trace plots, which indicated robust performance with no discernible trends in the final iterations (figures S3-5). Our final imputation models were validated and showed high agreement with the experimentally serotyped data, with an overall accuracy of 0·89 (0·85-0·92) for symptomatic infection and 0·78 (0·74-0·82) for inapparent infection (Table S3-6).

Throughout the study period, DENV serotypes 1, 2, and 3 were the dominant serotypes, representing 98% of total primary infections, reflecting the serotypes circulating in the country.^13^ In contrast, DENV4 circulation was very low and was exclusively detected in inapparent infections. The analysis of primary infections revealed distinct serotype distributions, with significant differences observed between the proportions of symptomatic versus inapparent infections caused by each serotype every year (figure 3A). For instance, in 2004, DENV1 was the only serotype detected in symptomatic cases, whereas all four serotypes circulated within the inapparent infection fraction. During the 2008-2010 period, DENV3 was dominant in symptomatic cases but exhibited lower relative prevalence within the inapparent fraction. Significant within-year variations in serotype distribution were observed between the symptomatic and inapparent infection fractions, particularly in 2004 (p 0·011), 2007 (p 0·021), 2008 (p 0·0025), and 2009 (p 0·005) (figure 3A, Table S7). These variations and temporal dynamics were consistent across experimentally serotyped and imputed datasets (figure 3A and figure S2C).

**Figure 3.**
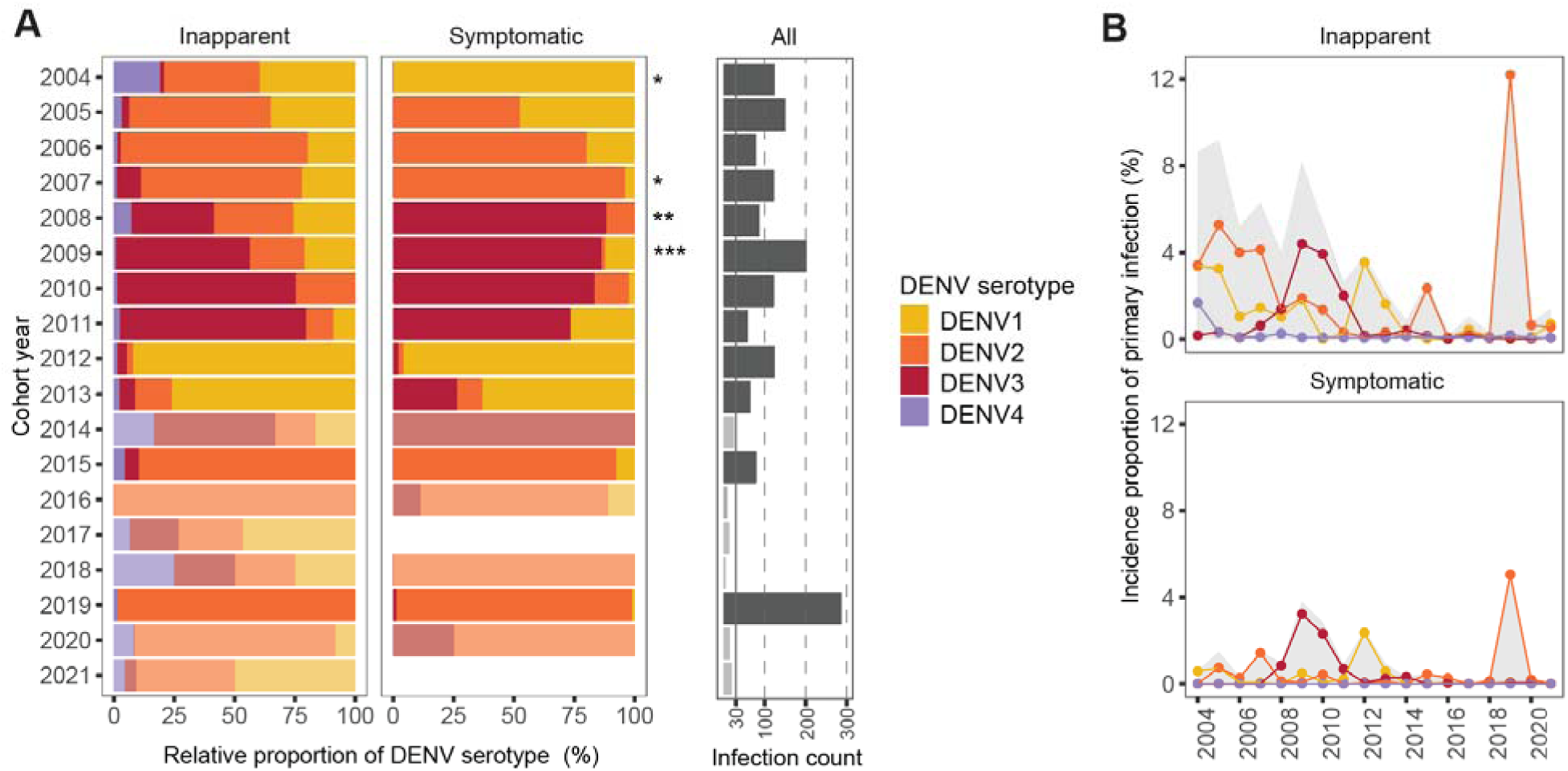
Serotype distribution in primary symptomatic and inapparent infections by serotype from 2004 to 2021 in the PDCS. **(A)** Pooled relative distribution of DENV serotypes circulating yearly in inapparent and symptomatic primary DENV infections across multiple imputations. Transparency was added in years with infection count <30. The gray panel shows the total incidence of primary DENV infections. **(B)** Temporal dynamics of the percent of primary DENV infections by infection outcome (shown in upper and lower panels) and serotype across imputations. The gray area covers the pooled incidence proportions of overall primary DENV infections by infection outcome. p-values: *, p<0·05; **, p<0·01; ***, p<0·001.

Subsequent stratification of the cumulative incidence proportions of inapparent and symptomatic infections by serotype (figure 3B) further accentuates the pattern observed initially: whereas symptomatic infections tend to be dominated by a single serotype, the DENV landscape unveiled by inapparent infections is more complex, with co-circulation of multiple DENV serotypes, including serotypes undetected in symptomatic surveillance, in multiple years.

DENV2 demonstrated the most extended circulation period and the highest overall incidence in our study compared to DENV1 and DENV3 (Table S8). Nevertheless, analyzing the cumulative annual incidence proportion of primary symptomatic and inapparent infections per serotype highlighted dynamic shifts in the dominance of DENV serotypes. While DENV2 dominated the initial years of the study, this dominance transitioned to DENV3 in 2009-2010 and subsequently shifted to DENV1 during the 2012-2013 period, before DENV2 resurgence in 2019 (figure 4A, Table S9). A marked change in DENV2 incidence proportion was particularly notable in 2019, with 17% (95% CI 15–19%) of the naïve population experiencing primary DENV2 infections compared to only 4% (95% CI 3–5%) in 2006 (figure 4A, figure S6, Table S9). The incidence proportion of primary DENV3 infections was particularly high between 2008 and 2011, reaching up to 7% (95% CI 6–9%) of the naïve population at its peak whereas for DENV1, incidence peaked in 2012 (6%, 95% CI 5–7%).

**Figure 4.**
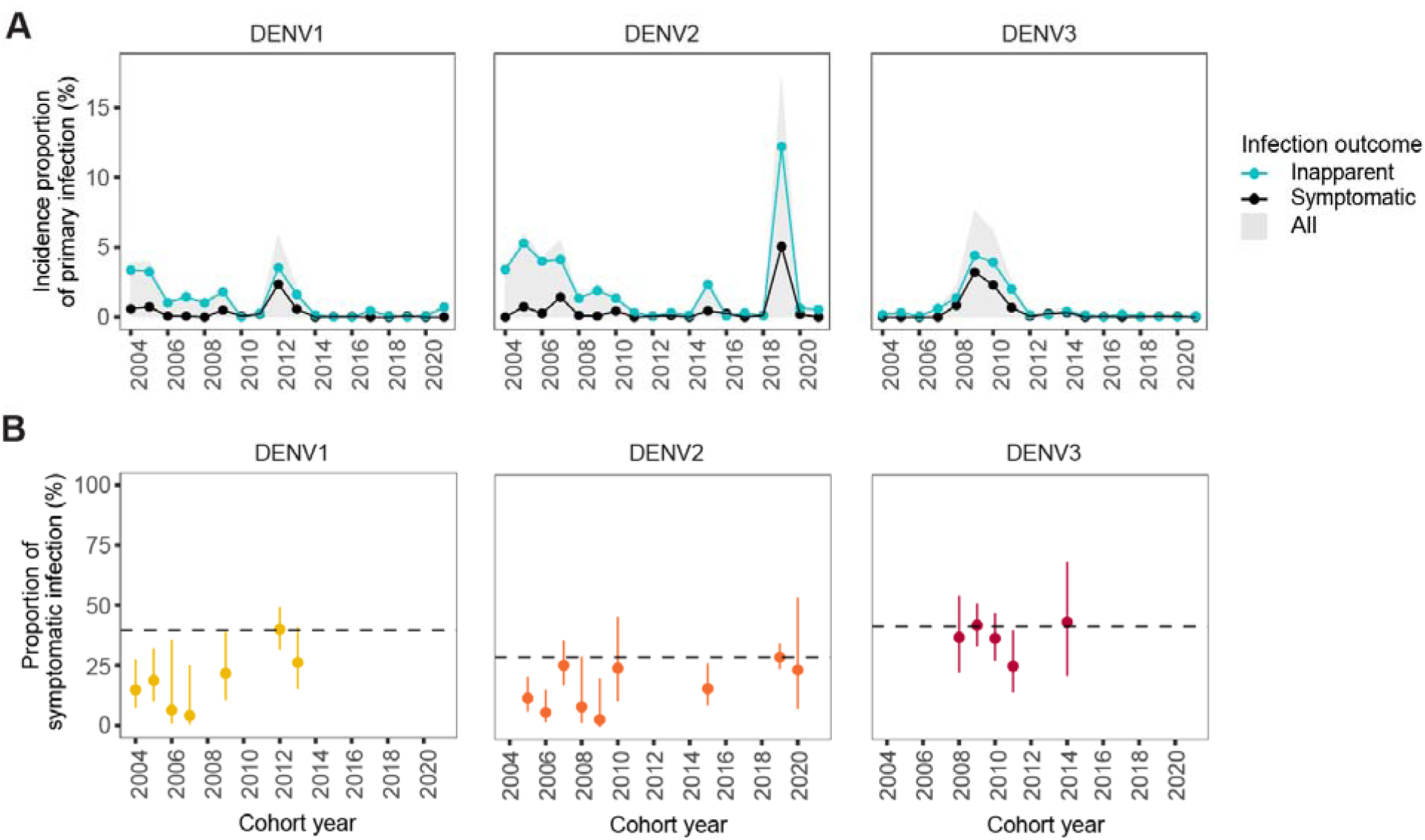
Temporal dynamics of symptomatic, inapparent, and overall primary DENV infections by serotype from 2004 to 2021 in the PDCS. **(A)** Pooled percent of primary DENV infections by infection outcome and serotype, as indicated in left, middle, and right panels across imputations. The gray area covers the pooled incidence proportions of overall primary DENV infections by serotype. **(B)** Proportion of symptomatic primary DENV infection by serotype (indicated in left, middle, and right panels). This analysis excludes years when no symptomatic infections or <10 primary infections were reported. This is the result of the pooled predicted marginal probabilities of disease given infection from logistic regressions adjusting for year, compared to reference years (dashed line) of the most intense outbreak (DENV1: 2012, DENV2: 2019, DENV3: 2009).

Analyzing the proportion of symptomatic infections among total infections per serotype over time revealed considerable yearly fluctuations. Excluding years when no symptomatic infections were found and years with <10 primary infections per serotype, the proportion of DENV1 symptomatic infections among DENV1 total infections ranged from 7% (95% CI 1–36%) in 2006 to a peak of 40% (95% CI 31%–49%) in 2012; DENV2 ranged from 6% (95% CI 2–16%) in 2006 to 29% (95%CI 24–35%) in 2019, and DENV3 ranged from 26% (95% CI 15–40%) in 2011 to 44% (95% CI 22–69%) in 2014 (figure 4B, Table S9). Notably, at the serotype level, years of the most intense outbreaks coincided with the highest proportion of symptomatic infections for all serotypes. To evaluate differences in virulence over time by serotype, we also report the values in pooled odds ratio form (Table S10-12).

Further investigation into the disease spectrum of primary infections underscored stark differences by serotype. Analyzing symptomatic versus inapparent and severe versus non-severe outcomes, DENV3 demonstrated significantly higher likelihood of symptomatic (Pooled Odds Ratio [POR] = 2·13, 95% CI 1·28–3·56) and severe infections (POR = 6·75, 95% CI 2·01–22·62), as evidenced by POR compared to DENV1 adjusting by epidemic year, whereas DENV2 did not show significant differences in either analysis compared to DENV1 (figure 5A-B). Such results were consistent across all the sensitivity analyses performed (Table S13).

**Figure 5.**
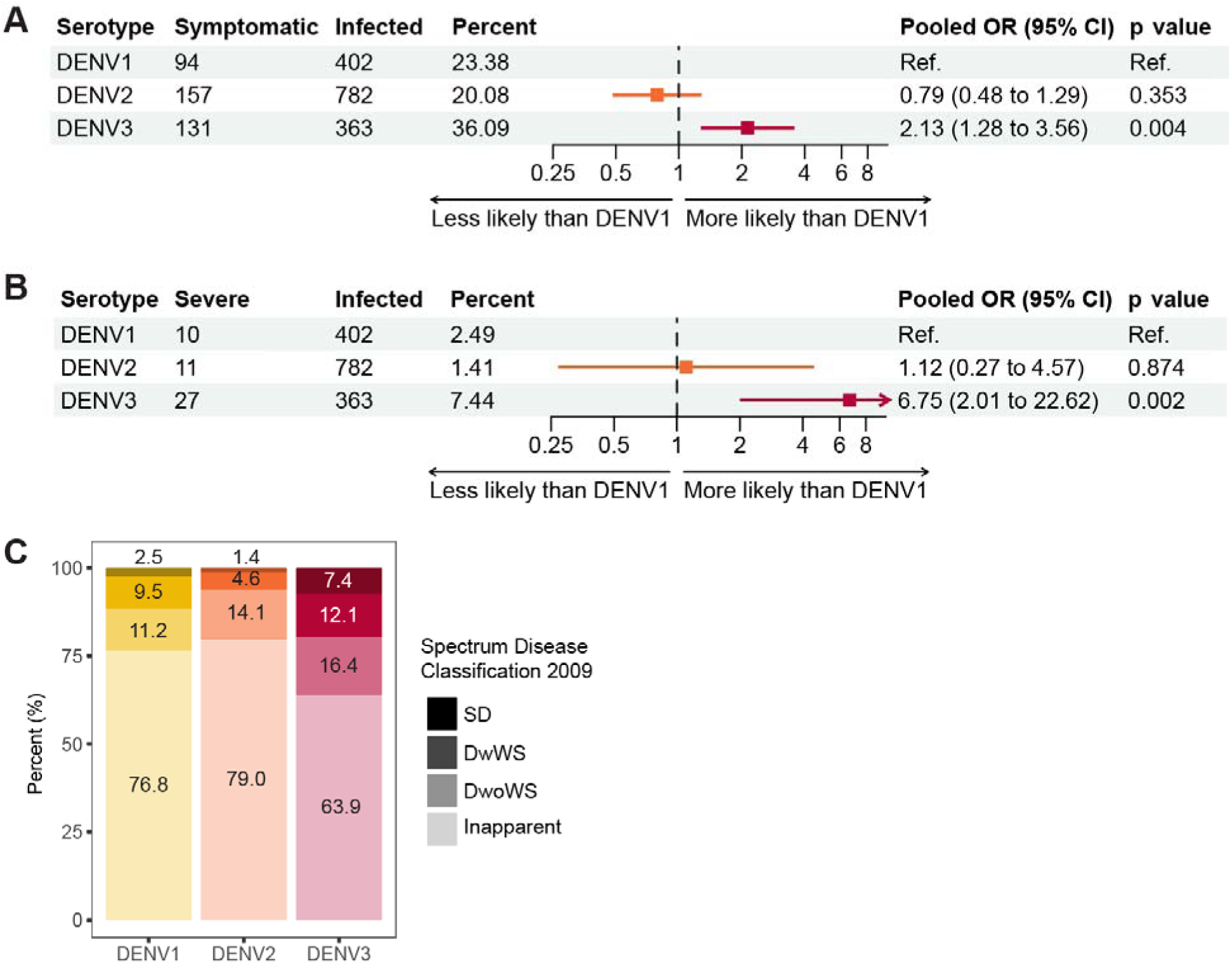
Spectrum of disease of primary DENV infections. (**A)** Pooled counts of symptomatic, overall, and crude percent of symptomatic primary DENV infections by DENV serotype, and pooled odds ratios (OR) adjusting by year showing the association between dengue case and serotype among primary DENV infections. **(B)** Pooled counts of severe, overall, and crude percent of severe primary DENV infections by DENV serotype, and pooled odds ratios (OR) adjusting by year showing association between dengue case and serotype among primary DENV infections. **(C)** Spectrum the of infection outcome and disease according to the 2009 classification as the crude pooled percentages of primary DENV infections.

Lastly, we summarized the spectrum of disease observed in primary infections caused by the three major DENV serotypes that circulated in Nicaragua between 2004 and 2021 by analyzing the frequency of inapparent, symptomatic, and severe DENV infections classified according to the WHO guidelines from 2009 (figure 5C). Although we found that a significant majority of primary infections remained inapparent, symptomatic infection was observed in 23·2% (19·2-27·8%), 20·1% (17·4-23·2%) and 36·1% (31·1-41·4%) of DENV1, DENV2, and DENV3 infection respectively. Among total primary infections, severe disease was observed in 2·5% (1·3-4·6%) of DENV1, 1·4% (0·8-2·6%) of DENV2, and 7·4% (5·1-10·6%) of DENV3 infections further highlighting the variability in infection outcomes associated with each serotype (Table S14).

## Discussion

Our study provides novel insights into serotype-specific epidemiological patterns and disease outcomes of primary DENV infections in Nicaragua by revealing the hidden contribution of inapparent infections. The EDIII multiplex microsphere-based assay has been previously evaluated to detect any prior ZIKV or prior DENV infection, showing a sensitivity of 94.9% and 100%, and a specificity of 97.8% and 98.5%, respectively.^11^ Here we expanded and evaluated its use for serotyping primary DENV infections, which demonstrated high performance when evaluated against the gold standard FRNT and RT-PCR assays. This significant methodological advancement offers a scalable and efficient alternative to the labor-intensive neutralization assays traditionally used to serotype infection. Compared to neutralization assays such as the FRNT, the microsphere-based assays are easier to standardize across laboratories and can be performed without the need for handling infectious materials under BSL2 conditions.

The majority of primary infections within our cohort were inapparent (76%), aligning with existing literature that suggests a high prevalence of inapparent and subclinical infection across dengue-endemic regions.^14–17^ However, the proportion of inapparent infections greatly fluctuates across years. The characterization of these infections allowed us to examine the serotype-specific proportion of primary inapparent infections and their temporal variability for the first time. Importantly, our study shows that while a single serotype often dominates symptomatic primary infections in any given period, the landscape of inapparent infections in Nicaragua is far more complex, revealing the co-circulation of multiple serotypes and serotype circulation undetected in symptomatic cases across multiple years. This observation suggests that the epidemiological footprint of DENV is broader and more nuanced than what is captured by monitoring symptomatic cases alone, whenever there is co-circulation of serotypes or hyperendemicity. This finding underscores the utility for continuous and comprehensive surveillance systems that capture both symptomatic and inapparent infections to accurately assess the epidemiological burden of DENV and inform public health interventions.

Temporal variations in the incidence and proportion of symptomatic primary infections further highlighted the evolving landscape of dengue in Nicaragua. Notably, we observed an increase in virulence of DENV1 over time, with more symptomatic primary infection in 2012 compared to 2004-2006. Furthermore, DENV2 infectivity surged in 2019, with 17% of the DENV-naïve population experiencing primary infections versus 5% in 2006. These changes could potentially be due to strain differences over time. Although our study does not directly link these observations to specific viral genetic variation, this hypothesis is supported by prior studies showing lineage shifts in DENV1 and DENV2 within our cohort (figure S7).^13, 18, 19^ Further, the level of population-level susceptibility to different serotypes, as well as changes in host or environmental factors may also influence infection dynamics and outcomes. For instance, the extensive circulation of ZIKV in 2016 may have enhanced the DENV2 epidemic in 2019.^5, 6, 20–22^ Additionally, increases in comorbidities (e.g., the rise in prevalence of obesity) in our cohort might also influence dengue disease burden.^23^ Finally, dengue cohort studies have reported large fluctuations in the yearly proportion of symptomatic infections and have determined that the predominant circulating serotype is one of the main factors driving this fluctuation.^8, 24^

By dissecting the full disease spectrum associated with primary DENV infection, our study revealed stark serotype-specific differences in infection outcome. Specifically, our finding supports and extends existing literature by demonstrating that DENV3 primary infections were not only significantly more severe,^25–27^ but also significantly more symptomatic compared to DENV1 and DENV2. For instance, a study of schoolchildren in Thailand found that schools with high DENV3 circulation were associated with lower detection of inapparent infections.^24^ Another study in Nicaragua found that among symptomatic infections, more severe primary infections are caused by DENV3 than by DENV1 or DENV2.^27^ Here, we expand on these results by taking the full spectrum of primary infections into consideration, including inapparent infections. Furthermore, our study shows that the low rate of symptomatic primary DENV2 infections in Nicaragua is not due to a scarcity of primary infections but rather due to a high rate of inapparent infections. Also, it is important to note that while severe dengue is more prevalent during secondary infections, it also occurs in primary infections, particularly with DENV3 and DENV1.^27–30^ Substantial morbidity and mortality can be attributed to primary infections, for example, ≥50% of severe cases and fatalities in a study in India^31^ and 23% of hospitalized dengue patients in a study in Mexico.^32^ Here, we also found a sizable amount of severe infections among primary cases, being the highest for DENV3. All together, these studies show that primary infections contribute significantly to disease burden. Thus, safe and effective vaccines for DENV-naïve individuals are warranted.

This study has several strengths. First, we expanded the use of a new simpler, high-throughput method for serotyping inapparent primary infections. Second, we analyzed 17 years of data based on a robust cohort study design that enables investigation of temporal dynamics of multiple DENV serotypes in a consistent, single setting with a large sample size. The study design captures inapparent, mild, and severe DENV infections, offering a comprehensive view of the disease burden that refines our understanding of DENV epidemiology. Further, while serotyping inapparent primary infections is not unique to our study, most studies are cross-sectional^33–35^ reporting “crude” seroprevalence of a given serotype. Here we report the evolution of the incidence of primary infection by serotype over time.

However, our study has some limitations, including minimal DENV4 circulation during the study timeframe. Nonetheless, among primary infections, we predict that DENV4 has a similar spectrum of symptomatic versus inapparent infection outcome compared to DENV2 as evidenced in the literature.^25^ The emergence of ZIKV might have impacted the DENV epidemiology in Nicaragua, which would affect our findings after 2016, especially for DENV2. Yet, when restricting our analyses from 2004-2015, our findings were similar. As our findings reflect the serotype distribution in Nicaragua, the generalizability of our findings may be somewhat limited by the regional focus and the genotype of local DENV strains (Table S15), which may have distinct virulence and infectivity. Nevertheless, we believe that our findings provide a robust rationale for future research in different settings to assess the epidemiology of other DENV genotypes and broader implications of our findings. The random selection of inapparent samples for EDIII serotyping likely mitigated selection bias, providing an unbiased estimate of serotype distribution among inapparent infections, but our imputation model assumes such data to be missing at random, dependent on time and space; deviations from this assumption could bias our findings. Our final imputation models were valid, except for DENV4, which accounted for only 2% of the inapparent infections. Our results remained robust when excluding DENV4 infections from our imputation models and across other sensitivity analyses. Finally, although we adjusted for age and sex in the imputation and main endpoint models, we cannot rule out residual confounding.

In summary, using a high throughput serotyping method to analyze inapparent primary DENV infections in a cohort over 17 years, our study enriches the epidemiological data landscape with unprecedented detail. Our findings reveal significant differences in the epidemiological profile of symptomatic and inapparent DENV infections and show serotype-specific patterns and disease spectra. The stark differences in disease outcome by serotype, particularly the symptomatic and severe nature of DENV3 infections versus others, highlight the need for balanced and effective vaccines across serotypes that provide protection even for those without prior DENV exposure. These insights are also important in refining existing models of DENV transmission and mark an advancement in our understanding of the serotype-specific risks of infection outcome, which allows us to better predict outbreaks and hospital admission, understand DENV transmission dynamics, and inform vaccine development.

## Supporting information

Transformed_EDIII-MMBA_Values

## Acknowledgements

We thank the study personnel at Centro de Salud Sócrates Flores Vivas, the Nicaraguan National Virology Laboratory, the Hospital Infantil Manuel de Jesús Rivera, and the Sustainable Sciences Institute. We are particularly grateful to the study participants and their families. This work was supported by the National Institute of Allergy and Infectious Diseases/National Institutes of Health (NIAID/NIH) via grants P01AI106695 (E.H.) and U01AI153416 (E.H.). The PDCS study was supported by NIAID/NIH grants P01AI106695 (E.H.), U01AI153416 (E.H.), U19AI118610 (E.H.), and R01AI099631 (A.B.), the Pediatric Dengue Vaccine Initiative grant VE-1 (E.H.) from the Bill and Melinda Gates Foundation, and NIH subcontract HHSN2722001000026C (E.H.).

## Authors contributions

Conceptualization: SB, JVZ, EH

Methodology: SB, JVZ, PL

Investigation: SB, JVZ, EMD, JH, ALG, CM

Visualization: SB, JVZ

Funding acquisition: AB, EH

Project administration: GK, AG, AB, EH

Manuscript writing – first draft: SB, JVZ, EH

Manuscript writing – review and editing: All authors

Access and data verification – SB, JVZ, EH

Manuscript submission decision - SB, JVZ, EH

## Data availability

All relevant data supporting the findings of this study are included within the manuscript or the supplementary appendix. Normalized EDIII-MMBA values from our evaluation set are accessible in the supplementary material “Transformed_EDIII-MMBA_Values.” We kindly request that researchers who utilize the data provided for their own studies acknowledge this by citing this paper.

Additional datasets generated and/or analyzed during the current study are available upon reasonable request, following the protocol approved by the Institutional Review Board (IRB) for the Pediatric Dengue Cohort Study. Researchers who wish to access the additional data are encouraged to submit a formal request to Dr. Eva Harris or the Committee for the Protection of Human Subjects at the University of California, Berkeley. In accordance with ethical guidelines and to ensure the proper use of the data, all requests will be reviewed and approved on a case-by-case basis. The associated code for this study is available for reference on Zenodo (doi: 10.5281/zenodo.12349676).

## References

1 Yang X, Quam MBM, Zhang T, Sang S. Global burden for dengue and the evolving pattern in the past 30 years. J Travel Med 2021; 28. DOI:10.1093/jtm/taab146.

2 Lee SY, Shih H-I, Lo W-C, Lu T-H, Chien Y-W. Discrepancies in dengue burden estimates: a comparative analysis of reported cases and global burden of disease study, 2010-2019. J Travel Med 2024; 31. DOI:10.1093/jtm/taae069.

3 Katzelnick LC, Gresh L, Halloran ME, et al. Antibody-dependent enhancement of severe dengue disease in humans. Science 2017; 358: 929–32.

4 Sabchareon A, Wallace D, Sirivichayakul C, et al. Protective efficacy of the recombinant, live-attenuated, CYD tetravalent dengue vaccine in Thai schoolchildren: a randomised, controlled phase 2b trial. Lancet 2012; 380: 1559–67.

5 Katzelnick LC, Narvaez C, Arguello S, et al. Zika virus infection enhances future risk of severe dengue disease. Science 2020; 369: 1123–8.

6 Zambrana JV, Hasund CM, Aogo RA, et al. Primary exposure to Zika virus is linked with increased risk of symptomatic dengue virus infection with serotypes 2, 3, and 4, but not 1. Sci Transl Med 2024; 16: eadn2199.

7 Bhoomiboonchoo P, Nisalak A, Chansatiporn N, et al. Sequential dengue virus infections detected in active and passive surveillance programs in Thailand, 1994-2010. BMC Public Health 2015; 15: 250.

8 Tsang TK, Ghebremariam SL, Gresh L, et al. Effects of infection history on dengue virus infection and pathogenicity. Nat Commun 2019; 10: 1246.

9 Ten Bosch QA, Clapham HE, Lambrechts L, et al. Contributions from the silent majority dominate dengue virus transmission. PLoS Pathog 2018; 14: e1006965.

10 Duong V, Lambrechts L, Paul RE, et al. Asymptomatic humans transmit dengue virus to mosquitoes. Proc Natl Acad Sci U S A 2015; 112: 14688–93.

11 Dahora L, Castillo IN, Medina FA, et al. Flavivirus Serologic Surveillance: Multiplex Sample-Sparing Assay for Detecting Type-Specific Antibodies to Zika and Dengue Viruses. 2023; published online April 7. DOI:10.2139/ssrn.4411430.

12 Kuan G, Gordon A, Avilés W, et al. The Nicaraguan pediatric dengue cohort study: study design, methods, use of information technology, and extension to other infectious diseases. Am J Epidemiol 2009; 170: 120–9.

13 Cerpas C, Vásquez G, Moreira H, et al. Introduction of New Dengue Virus Lineages of Multiple Serotypes after COVID-19 Pandemic, Nicaragua, 2022. Emerg Infect Dis 2024; 30: 1203–13.

14 Endy TP, Chunsuttiwat S, Nisalak A, et al. Epidemiology of inapparent and symptomatic acute dengue virus infection: a prospective study of primary school children in Kamphaeng Phet, Thailand. Am J Epidemiol 2002; 156: 40–51.

15 Morrison AC, Minnick SL, Rocha C, et al. Epidemiology of dengue virus in Iquitos, Peru 1999 to 2005: interepidemic and epidemic patterns of transmission. PLoS Negl Trop Dis 2010; 4: e670.

16 Asish PR, Dasgupta S, Rachel G, Bagepally BS, Girish Kumar CP. Global prevalence of asymptomatic dengue infections - a systematic review and meta-analysis. Int J Infect Dis 2023; 134: 292–8.

17 Tissera H, Amarasinghe A, De Silva AD, et al. Burden of dengue infection and disease in a pediatric cohort in urban Sri Lanka. Am J Trop Med Hyg 2014; 91: 132–7.

18 Thongsripong P, Edgerton SV, Bos S, et al. Phylodynamics of dengue virus 2 in Nicaragua leading up to the 2019 epidemic reveals a role for lineage turnover. BMC Ecol Evol 2023; 23: 58.

19 Edgerton SV, Thongsripong P, Wang C, et al. Evolution and epidemiologic dynamics of dengue virus in Nicaragua during the emergence of chikungunya and Zika viruses. Infect Genet Evol 2021; 92: 104680.

20 Ribeiro GS, Hamer GL, Diallo M, Kitron U, Ko AI, Weaver SC. Influence of herd immunity in the cyclical nature of arboviruses. Curr Opin Virol 2020; 40: 1–10.

21 Pinotti F, Giovanetti M, de Lima MM, et al. Shifting patterns of dengue three years after Zika virus emergence in Brazil. Nat Commun 2024; 15: 632.

22 Estofolete CF, Versiani AF, Dourado FS, et al. Influence of previous Zika virus infection on acute dengue episode. PLoS Negl Trop Dis 2023; 17: e0011710.

23 Mercado-Hernandez R, Myers R, Carillo FB, et al. Obesity is associated with increased pediatric dengue virus infection and disease: A 9-year cohort study in Managua, Nicaragua. medRxiv. 2024; DOI: 2024.04.02.24305219.

24 Endy TP, Anderson KB, Nisalak A, et al. Determinants of inapparent and symptomatic dengue infection in a prospective study of primary school children in Kamphaeng Phet, Thailand. PLoS Negl Trop Dis 2011; 5: e975.

25 Soo K-M, Khalid B, Ching S-M, Chee H-Y. Meta-Analysis of Dengue Severity during Infection by Different Dengue Virus Serotypes in Primary and Secondary Infections. PLoS One 2016; 11: e0154760.

26 Cordeiro MT, Silva AM, Brito CAA, et al. Characterization of a dengue patient cohort in Recife, Brazil. Am J Trop Med Hyg 2007; 77: 1128–34.

27 Narvaez F, Montenegro C, Juarez JG, et al. Dengue severity by serotype in 19 years of pediatric clinical studies in Nicaragua. medRxiv 2024; published online Feb 13. DOI:10.1101/2024.02.11.24302393.

28 Nisalak A, Endy TP, Nimmannitya S, et al. Serotype-specific dengue virus circulation and dengue disease in Bangkok, Thailand from 1973 to 1999. Am J Trop Med Hyg 2003; 68: 191–202.

29 Balmaseda A, Hammond SN, Pérez L, et al. Serotype-specific differences in clinical manifestations of dengue. Am J Trop Med Hyg 2006; 74: 449–56.

30 Harris E, Videa E, Pérez L, et al. Clinical, epidemiologic, and virologic features of dengue in the 1998 epidemic in Nicaragua. Am J Trop Med Hyg 2000; 63: 5–11.

31 Aggarwal C, Ahmed H, Sharma P, et al. Severe disease during both primary and secondary dengue virus infections in pediatric populations. Nat Med 2024; published online Feb 6. DOI:10.1038/s41591-024-02798-x.

32 Annan E, Treviño J, Zhao B, Rodriguez-Morales AJ, Haque U. Direct and indirect effects of age on dengue severity: The mediating role of secondary infection. PLoS Negl Trop Dis 2023; 17: e0011537.

33 Sasmono RT, Taurel A-F, Prayitno A, et al. Dengue virus serotype distribution based on serological evidence in pediatric urban population in Indonesia. PLoS Negl Trop Dis 2018; 12: e0006616.

34 Alagarasu K, Tomar S, Patil J, et al. Seroprevalence of dengue virus infection in Pune City in India, 2019: A decadal change. J Infect Public Health 2023; 16: 1830– 6.

35 Limothai U, Tachaboon S, Dinhuzen J, et al. Dengue pre-vaccination screening test evaluation for the use of dengue vaccine in an endemic area. PLoS One 2021; 16: e0257182.

