## Supplementary appendix for "Serotype-Specific Epidemiological Patterns of Inapparent versus Symptomatic Primary Dengue Virus Infections: A 17-year cohort study in Nicaragua"

### Table of contents


**Supplementary Methods** **2**
Ethics statement 2
The Pediatric Dengue Cohort Study and Participants 2
Case and inapparent infection definition 2
Detection of cases 3
Inapparent infections 3
Severity classification as per the 2009 WHO classification 3
EDIII-MMBA 3
Neutralization assay 4
DENV iELISA 4
Typing of primary inapparent infection based on EDIII-MMBA 5
Evaluation of the EDIII-MMBA 5
Covariate definition and relationship between variables 6
Sensitivity analyses 6
Evaluation of the final imputation models 7
**Supplementary figures and tables** **8**
Table S1. Count of naïve participants and primary DENV infections per year 8
Table S2. Breakdown of serotyped and non-serotyped inapparent and symptomatic primary DENV infections 9
Table S3. Imputation model performance on inapparent infections based on the experimentally serotyped data 10
Table S4. Imputation model performance on symptomatic infections based on the experimentally serotyped data 10
Table S5. Actual vs predicted inapparent DENV infections based only on the experimentally serptyped data 11
Table S6. Actual vs predicted symptomatic DENV infections based only on the experimentally serotyped data 11
Table S7. Significance of fisher tests 12
Table S8. Overall incidence of primary DENV infections by serotype. 13
Table S9. Risk of dengue infections and percent symptomatic given primary infection by dengue serotype over time, based on model predictions across imputations excluding years with low symptomatic circulation (n < 10). 13
Table S10. Odds ratios of symptomatic primary infection given DENV1 infection over time 14
Table S11. Odds ratios of symptomatic primary infection given DENV2 infection over time 14
Table S12. Odds ratios of symptomatic primary infection given DENV3 infection over time 14
Table S13. Sensitivity analyses 15
Table S14. Outcome of primary infections 17
Table S15. Genotype of Nicaraguan DENV serotypes. 18
Figure S1. Flowchart of the screening and serotyping process to study the epidemiology of primary DENV infections in the PDCS 19
Figure S2. Distribution of serotyped, non-serotyped, and imputed symptomatic and inapparent primary DENV from 2004 to 2021 in the PDCS 20
Figure S3. Trace plots for the final imputation models 21

Figure S4. Trace plots for the final imputation framework based on only 10 simulations 22
Figure S5. Trace plots for the final imputation framework based on only 10 simulations and 30 iterations 23
Figure S6. Temporal dynamics of overall primary DENV infections by serotype from 2004 to 2021 in the PDCS. 24
Figure S7. Temporal shift in Nicaraguan DENV2 lineages 25
**Supplementary References 26**

### Supplementary Methods

#### Ethics statement

The human subjects protocol for the Pediatric Dengue Cohort Study (PDCS) was reviewed and approved by the Institutional Review Boards (IRB) of the University of California, Berkeley (2010-09-2245), the University of Michigan (HUM00091606), and the Nicaraguan Ministry of Health (CIRE-09/03/07-008). Parents or legal guardians of all subjects provided written informed consent, and subjects 6 years of age and older provided assent.

#### The Pediatric Dengue Cohort Study and Participants

The PDCS is an ongoing prospective study initiated in 2004 and carried out at the Sócrates Flores Vivas Health Center (HCSFV), located in District 2 of Managua, Nicaragua.[1] Located adjacent to Lake Xoxitlan, District 2 encompasses neighborhoods spanning a spectrum of low to middle socioeconomic status. The HCSFV serves ~62,000 residents across 18 neighborhoods within District 2.[2] The illiteracy rate in this area is approximately 7%, and over 90% of the households have access to running water and a sewage system. All residents have access to electricity.[3]

The PDCS tracks approximately 4,000 active participants yearly and has followed ~9,800 children since its inception. To preserve the original age distribution of the cohort, approximately 300 two-year-old children, along with children aged 3-11 years as needed, are enrolled annually. Noteworthy adjustments to the age limit were implemented in 2018, 2019, and 2022, extending the eligibility to 15, 16, and 17 years, respectively. Participants benefit from free, round-the-clock medical care provided by study physicians at the HCSFV. In cases requiring hospitalization, study staff facilitate patient transfer to the study hospital, the National Pediatric Reference Hospital (Hospital Infantil Manuel de Jesús Rivera). Children presenting with fever and since July 2016, rash, are screened for signs and symptoms of dengue, Zika, and chikungunya. Symptomatic infections are captured through passive surveillance, complemented with periodic home visits. For each suspected case, both acute (0-5 days post-symptom onset) and convalescent samples (14-21 days) are collected to confirm infection. Approximately 97% of cases present to the HCSFV within the first three days of illness, and convalescent samples are collected from 94% of cases.[1] Additionally, the study collects annual healthy blood samples from all participants in March-April (previously in July before 2011), which enable the detection of inapparent arboviral infections.[4]

A series of annual surveys are administered verbally, and answers are recorded on smartphones or tablet computers using custom-built software. Two questionnaires are administered, one for every household and one for every participant. The participant questionnaire covers demographic information such as biological sex (with no genetic confirmation), age, and education level. The household questionnaire includes questions on assets and conditions of the home.

#### Case and inapparent infection definition

Upon medical examination, children enrolled in the study are categorized into A, B, C, or D cases, as follows: A: individuals meeting the 1997 or 2009 World Health Organization (WHO) case definition for dengue; B: those with undifferentiated fever of undefined origin; C: children with an acute febrile illness identified as a disease other than dengue, chikungunya, or Zika; D: individuals with other conditions (including rash) that do not include fever. A comprehensive array of clinical and laboratory information, encompassing 189 different variables4 is collected for each participant. This includes vital signs, temperature, musculoskeletal pain, respiratory symptoms, gastrointestinal symptoms, indicators of dehydration, rashes, and other skin anomalies. Cases categorized as A and B undergo testing for acute

DENV and, since 2014, chikungunya virus (CHIKV) infection. Since 2015, cases meeting the criteria of A, B, or D (rash without fever) are also tested for acute Zika virus (ZIKV) infection.

*Detection of cases*

Acute samples are examined using the ZCD multiplex real-time reverse transcriptase-PCR (rRT-PCR) assay for DENV, ZIKV, and CHIKV;[5] before the introduction of ZIKV in Nicaragua, different RT-PCR methods were used.[6,7] DENV-positive samples are then serotyped by multiplex rRT-PCR, and a subset undergo virus isolation in C6/36 cells.[7,8] Evaluation of paired acute-phase and convalescent-phase samples is conducted to detect seroconversion of IgM antibodies, indicating current infection. This is done using separate in-house DENV and ZIKV IgM capture (MAC)-ELISA assays.[9,10] DENV inhibition ELISA (iELISA) and ZIKV iELISA assays are used to measure total anti-DENV and -ZIKV antibodies, respectively, and to detect seroconversion or a ≥4-fold increase in antibody titer. Additionally, a computer algorithm assists in identifying Zika cases from dengue cases based on the results of the aforementioned serological tests.[11] Those with serotyping by RT-PCR that was inconclusive due to unclear results or low viremia, which were positive by either MAC ELISA or iELISA were considered DENV serology cases.

*Inapparent infections*

Before 2016, inapparent DENV infections were identified if a seroconversion or a ≥4-fold increase in antibody titer between annual samples was observed, as measured by DENV iELISA, in the absence of any observed febrile episode identified as a dengue case in the intervening year.[12] Following the 2016 Zika epidemic, inapparent DENV infections were defined as seroconversion or a ≥4-fold rise in DENV iELISA titer in absence of ZIKV seroconversion (measured by ZIKV NS1 BOB ELISA or ZIKV iELISA) or an observed dengue, Zika, or flavivirus case in that year.[9] Seroconversion by ZIKV NS1 BOB ELISA and/or seroconversion or >4-fold rise by ZIKV iELISA was used to detect inapparent ZIKV infections.

**Severity classification as per the 2009 WHO classification**

*Dengue without Warning Signs (DwoWS):* A laboratory-confirmed dengue case manifesting fever and two of the following clinical features: nausea/vomiting, rash, aches and pains, positive tourniquet test, leukopenia.

*Dengue with Warning Signs (DwWS):* A laboratory-confirmed dengue case manifesting symptoms such as abdominal pain, persistent vomiting, fluid accumulation, lethargy, mucosal bleeding, liver enlargement, or a rise in hematocrit concurrent with a decrease in platelet count.[13]

*Severe Dengue (SD):* A laboratory-confirmed dengue case featuring severe bleeding, significant plasma leakage leading to shock, fluid accumulation resulting in respiratory distress, and/or organ failure. Organ involvement was indicated by liver ALT or AST levels of ≥1,000, impaired consciousness, or heart or other organ failures.

**EDIII-MMBA**

Recombinant envelope domain III (EDIII) proteins of DENV1-4 and ZIKV (gift from Lakshmanane Premkumar, University of North Carolina, Chapel Hill) were site-specifically biotinylated, conjugated to avidin-coated MagPlex Luminex beads and combined. Immune complexes were formed in 384-well plates by mixing appropriately diluted plasma (four 4-fold dilutions starting at 1:50) with antigen-coupled microspheres (500 microsphere/antigen/well) for 90 minutes at 37°C, shaking at 1200 rpm. After incubation, plates were washed using an automatic magnetic washer (Tecan Hydrospeed) with phosphate-buffered saline (PBS) containing 0.1% bovine serum albumin (BSA) and 0.02% Tween 20. Antigen-specific IgG binding was detected using phycoerythrin (PE)-coupled mouse anti-human total IgG (SouthernBiotech). Median fluorescence intensity (MFI) was measured using an iQue3 (Intellicyt) instrument set to acquire at least 50 beads per bead region. DENV-naïve human serum (NHS) was run in triplicate on each plate. Positive signal was defined as an MFI greater than the average MFI of NHS + 3 SD at the highest dilution (1:50). Negative signal (e.g., absence of EDIII antibody) was defined as an MFI lower than the average MFI of NHS + 3 SD at the highest dilution (1:50).

**Neutralization assay**

The focus reduction neutralization test (FRNT) assay was conducted using Vero cells and clinical isolates from Nicaragua (DENV1 5575.10a1SPD1, DENV2 8891.12a1SPD2, DENV3 6629.10a1SPD3, and ZIKV Nica2-16) or DENV4 Sri Lanka 92 strain (NCBI KJ160504.1) as previously described.[14] Briefly, cells were seeded in a 96-well plate one day before infection. Serum/plasma were serially diluted and mixed with an equal inoculum of DENV at a volume ratio of 1:1 to form immune complexes. The virus-antibody mixture was incubated for 1 hour at 37°C in 5% $CO_2$. Cell substrate growth medium was removed, and the virus-antibody mixture was added to the cells and incubated for 1 hour at 37°C in 5% $CO_2$. Subsequently, a 0.6% carboxymethylcellulose overlay was added, and the plates were incubated for 48 hours. Foci were developed and counted using an CTL Immunospot analyzer (Cellular Technology, Ltd.). Foci counts were fitted using a variable slope dose response curve using Graphpad, and neutralizing antibody titer ($NT_{50}$) values were calculated with constrained top and bottom values of 100 and 0, respectively. Percent relative infection was calculated as a ratio of foci counts in each serial dilution to the foci counts from the final serial dilution of each sample. All samples and mAbs were run in duplicate with reported values required to have an $R^2$ > 0.85 and a Hill slope > 0.5. The serotype responsible for the inapparent infection was identified as the serotype to which the serum showed the highest neutralization.

**DENV iELISA**

The inhibition ELISA (iELISA) protocol,[12] adapted from the method by Fernández and Vázquez[15], is a diagnostic assay to detect antibodies against DENV in serum samples. The protocol involves coating 96-well polystyrene plates with anti-DENV polyclonal human IgG and incubating overnight at room temperature. After washing, the wells are blocked with BSA, then incubated at 37°C for 30 minutes. Next, DENV1-4 antigen mix is added to the wells, and the plates are again incubated, followed by washing. Serum samples, diluted according to the patient's DENV history as either a single 1:10 dilution (if the child was DENV-negative in the previous years) or four 10-fold dilutions (if the child is DENV-immune), are then added and incubated. A negative control (normal human serum) and a positive control (serum with high DENV-specific antibody titer) are also included on the plate. Following another incubation period and washing, a horse-radish peroxidase (HRP)-conjugated secondary antibody is added. After a final incubation and wash, the colorimetric peroxidase substrate TMB is introduced, and the plates are protected from light during a room temperature incubation of 30 minutes. The reaction is stopped with sulfuric acid, and absorbance is read at 450/630nm. The percentage of inhibition for each sample is determined relative to the negative control's absorbance, considering 100% as the mean negative control absorbance.

Seroconversion is identified when absorbance at a 1:10 dilution is less than 50% in the first year but equal to or more than 50% in the subsequent year. If percent inhibition is under 50% at the 1:10 dilution, the titer is reported as <1:10. For those who have seroconverted previously, titration of serial dilutions enables estimation of the iELISA titer using the Reed-Muench method[16], which accounts for the dilution factor and the percent inhibition before and after the 50% inhibition threshold.

**Typing of primary inapparent infection based on EDIII-MMBA**

During the evaluation phase, the serotype was assigned based on the highest MFI value observed among the set of EDIII antigens. In order to define more stringent rules to serotype an inapparent infection in the larger cohort set, we analyzed the pattern of cross-reactivity observed in the evaluation set using the MFI values of the EDIII set at a serum dilution of 1:800, chosen because this dilution was the common dilution outside of the prozone for all samples in the evaluation set. Specifically, the MFI values for EDIII-DENV1, EDIII-DENV2, EDIII-DENV3, EDIII-DENV4, and EDIII-ZIKV were expressed as the relative proportion of the highest MFI observed. We generated a heatmap (example shown in Fig. 2) with hierarchical clustering by Wald’s method, which resulted in 5 distinct clusters separating DENV1-4 and ZIKV with accuracy, also providing visual support for analyzing cross-reactivity patterns (examples shown in Fig. 2). Based on this analysis, we established empirical cut-off values for each serotype based on the relative MFI observed in the evaluation set by comparing the relative proportion of the second-highest MFI to the highest MFI observed. Specifically, the cut-off values applied derived from the evaluation set applied to the cohort analysis were as follows: second highest MFI observed less than 70% of the DENV1 signal in DENV1 primary infection, less than 30% of the DENV2 signal in DENV2 primary infection, less than 60% of the DENV3 signal in DENV3 primary infection, less than 40% of the DENV4 signal in DENV4 primary infection, and less than 20% of the ZIKV signal in ZIKV primary infection.

Next, the EDIII-MMBA was applied to the larger set of the PDCS, and serotyping was performed according to two criteria: (A) the serotype was assigned based on the highest MFI value observed among a set of EDIII antigens, and (B) the validity of the serotyping was assessed according to the empirical cutoff mentioned above. Serotyping was considered valid when the second highest relative MFI value was below the empirical cutoff value of the respective maximum MFI. For example, if the maximum MFI indicated a DENV1 serotype, the serotyping was valid if the second highest MFI was below 70%; it was invalid if the second highest MFI was above 70%.

**Evaluation of the EDIII-MMBA**

The evaluation of the EDIII-MMBA for serotyping primary inapparent infection involved two phases, both using samples from the PDCS: (1) evaluation using samples from participants with symptomatic primary DENV infections, and (2) evaluation using samples from participants with primary inapparent DENV infections. Samples were selected by convenience, which were readily available at UC Berkeley with sufficient volume, infection history data showing naïve DENV status by DENV iELISA prior to the infection event, RT-PCR results in the case of symptomatic DENV infections and neutralization data in the case of inapparent infections. The EDIII MMBA data was generated after sample selection, and samples were processed blindly. The results obtained were compared against the gold standard serotyping results conducted by an independent researcher. Of note, DENV4 circulated at epidemic levels in 2022-2023, right after our study ended. We then had the opportunity to include several DENV4 samples from this epidemic to evaluate our EDIII-MMBA assay against DENV4 primary infection serotyped by RT-PCR.

Phase I

Serum samples were selected from 70 participants with symptomatic primary DENV infections (DENV1=16, DENV2=13, DENV3=20, DENV4=6, and ZIKV=15), confirmed by gold-standard realtime RT-PCR.

Phase II

Serum samples were selected from 86 participants with primary inapparent DENV infections (DENV1=38, DENV2=19, DENV3=19, and ZIKV=10), confirmed by gold-standard antibody focus reduction neutralization test."

*Performance evaluation*

The accuracy of the EDIII-MMBA for serotype identification in primary DENV infections was evaluated by comparing it to gold standards: RT-PCR and FRNT assays. Sensitivity, specificity, and accuracy, the latter calculated as the sum of the true positives and true negatives divided by total number of samples evaluated, were calculated for each DENV serotype and Zika virus (ZIKV) across all test combinations. We calculated 95% confidence intervals using the exact binomial method for all the performance measures.

**Covariate definition and relationship between variables**

Covariates in our study include: DENV serotype, epidemic year, neighborhood and household location.

*DENV serotype*: DENV serotype determined by RT-PCR or EDIII-MMBA in symptomatic primary DENV infections or inapparent primary DENV infections, respectively. This exposure variable was analyzed as a categorical variable.

*Epidemic year*: The epidemic year starts between February-April (in July before 2011), corresponding to the start of the annual sampling, and ends at the beginning of the next annual sampling. During the annual sampling this period, the season is dry and DENV circulation is low. Cases usually peak between September and October during the wet season. This covariate was analyzed as a categorical variable.

*Neighborhood:* Neighborhood was defined by political administration by the Nicaraguan Census where participants live. This covariate was analyzed as a categorical variable.

Variable relationship: A causal framework for the main endpoint analysis (e.g. ratio of symptomatic versus inapparent and severe versus non-severe DENV infections by serotype) was created using Directed Acyclic Graph (DAG). Yellow shade shows exposure variables while pink shade shows the outcome variable. White boxes show adjusted/controlled variables. Black lines show closed paths, and the green lines show unbiased open path. The dashed line shows the presumptive causal relationship. The analyses were restricted to individuals with naïve status and adjusted by year. Age was adjusted in both the imputation models and main endpoint models and are shown as sensitivity analyses.

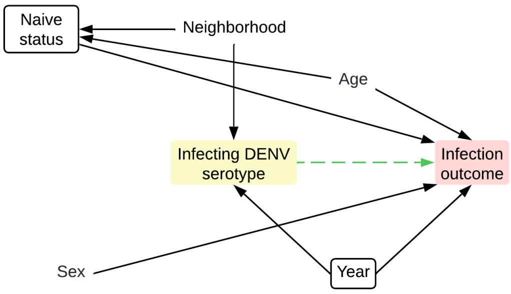


**Sensitivity analyses**

We performed several sensitivity analyses on our main endpoint models, including: (1) logistic regression analysis on the experimentally serotyped data, excluding missing data; (2) addition of the "no EDIII-Abs detected" category into the polytomous regression to address the potential limitation of the EDIII-MMBA to serotype primary infections detected by iELISA; (3) performing imputations with various frameworks, (3) including the non-parametric models (not assuming a specific form for the relationship between predictors and the response) such as regression trees, which partition the data into subsets based on feature values, creating a tree-like model for predictions being easy to interpret but prone to overfitting; (4) random forest, which is an ensemble method that combines the predictions from multiple decision trees to make more accurate and stable estimations being more robust to overfitting due to the averaging of multiple trees; (5) the semi-parametric predictive mean matching framework, which retains the original distribution of the data assuming that the distribution of the missing data is similar to the observed data; (6) adjusting the endpoint models not only by year but also by age and biological sex (no genetic confirmation); (7) changing the covariates of our imputation models to include only year; (8) adding year with an interaction term with region (neighborhoods classified by northeast, northwest, southeast, and southwest); (9) incorporating year with an interaction term with region plus age; (10) ignoring DENV4 cases and imputing only DENV1-3 cases; (11) restricting our analyses from 2004 to 2015 to only cover the pre-ZIKV era; and (12) removing the 76 samples from the EDIII-MMBA validation, which were selected by convenience sampling. All imputation models to address missing data were stratified by inapparent and symptomatic infections and all main endpoint models were adjusted by year.

We tested multiple frameworks to ensure that our analyses were not compromised by incorrect or overly restrictive assumptions or by the selection of models susceptible to bias or variance. For instance, simpler models such as polytomous regression often exhibit high bias with low variance, whereas more complex models, like random forests, typically exhibit low bias yet high variance. Finally, we assessed models based on a trade-off between interpretability and accuracy. While polytomous regression models are highly interpretable (the optimal), random forests, despite their accuracy, tend to be less so.

**Evaluation of the final imputation models**

We first examined trace plots to assess the convergence of our final imputation models, which adjust for year and neighborhood, constructed using 1000 simulations and 10 iterations. To further scrutinize visually the convergence and potential trends within the models on a more granular level, we performed additional runs with reduced simulations (10 simulations) across various numbers of iterations (10 and 30) to ensure stability across iterations. Additionally, we compared the model outputs to experimentally serotyped data, focusing on metrics such as overall accuracy, kappa statistics, and serotype-specific sensitivity and specificity. Lastly, we assured that the models were effective in estimating the distribution of the number of individuals by serotype and year, which was essential as our primary goal was to infer statistical parameters rather than individual imputations.

### Supplementary figures and tables

#### Table S1. Count of naïve participants and primary DENV infections per year

| | Naive | Infection | | Inapparent | | Symptomatic | | | | | |
|---|---|---|---|---|---|---|---|---|---|---|---|
| | | | | | | | | DENV1 | DENV2 | DENV3 | Serology[1] |
| | n | n | % | n | % | n | % | n | n | n | n |
| 2004 | 1289 | 124 | 9·62 | 116 | 93·55 | 8 | 6·45 | 6 | 0 | 0 | 2 |
| 2005 | 1328 | 150 | 11·3 | 129 | 86 | 21 | 14·00 | 10 | 10 | 0 | 1 |
| 2006 | 1406 | 79 | 5·62 | 74 | 93·67 | 5 | 6·33 | 1 | 4 | 0 | 0 |
| 2007 | 1511 | 123 | 8·14 | 99 | 80·49 | 24 | 19·51 | 1 | 22 | 0 | 1 |
| 2008 | 1713 | 87 | 5·08 | 70 | 80·46 | 17 | 19·54 | 0 | 2 | 14 | 1 |
| 2009 | 1595 | 202 | 12·66 | 137 | 67·82 | 65 | 32·18 | 8 | 1 | 53 | 3 |
| 2010 | 1433 | 123 | 8·58 | 80 | 65·04 | 43 | 34·96 | 0 | 5 | 32 | 6 |
| 2011 | 1624 | 59 | 3·63 | 44 | 74·58 | 15 | 25·42 | 3 | 0 | 10 | 2 |
| 2012 | 1861 | 124 | 6·66 | 75 | 60·48 | 49 | 39·52 | 44 | 1 | 1 | 3 |
| 2013 | 2082 | 65 | 3·12 | 45 | 69·23 | 20 | 30·77 | 11 | 2 | 4 | 3 |
| 2014 | 2115 | 25 | 1·18 | 18 | 72 | 7 | 28·00 | 0 | 0 | 1 | 6 |
| 2015 | 2543 | 80 | 3·15 | 68 | 85 | 12 | 15·00 | 0 | 8 | 0 | 4 |
| 2016 | 2556 | 9 | 0·35 | 2 | 22·22 | 7 | 77·78 | 0 | 3 | 0 | 4 |
| 2017 | 1425 | 14 | 0·98 | 14 | 100 | 0 | 0·00 | 0 | 0 | 0 | 0 |
| 2018 | 1492 | 5 | 0·34 | 3 | 60 | 2 | 40·00 | 0 | 1 | 0 | 1 |
| 2019 | 1356 | 287 | 21·17 | 203 | 70·73 | 84 | 29·27 | 0 | 47 | 0 | 37 |
| 2020 | 1459 | 15 | 1·03 | 12 | 80 | 3 | 20·00 | 0 | 1 | 0 | 2 |
| 2021 | 1561 | 20 | 1·28 | 20 | 100 | 0 | 0·00 | 0 | 0 | 0 | 0 |

[1] Positive according to serological assays (MAC-ELISA, or iELISA), with inconclusive RT-PCR results.

**Table S2. Breakdown of serotyped and non-serotyped inapparent and symptomatic primary DENV infections**

| Cohort year | Inapparent | | | | Symptomatic | | |
|---|---|---|---|---|---|---|---|
| | Overall n (%) | Screened | Typed[1] n (%) | Non-screened[2] n (%) | Overall n (%) | Typed[1] n (%) | Non-typed[2] n (%) |
| 2004 | 116 (100) | 46 (40) | 40 (34) | 76 (66) | 8 (100%) | 6 (75%) | 2 (25%) |
| 2005 | 129 (100) | 50 (39) | 39 (30) | 90 (70) | 21 (100%) | 20 (95%) | 1 (4·8%) |
| 2006 | 74 (100) | 30 (41) | 13 (18) | 61 (82) | 5 (100%) | 5 (100%) | 0 (0%) |
| 2007 | 99 (100) | 38 (38) | 23 (23) | 76 (77) | 24 (100%) | 23 (96%) | 1 (4·2%) |
| 2008 | 70 (100) | 27 (39) | 14 (20) | 56 (80) | 17 (100%) | 16 (94%) | 1 (5·9%) |
| 2009 | 137 (100) | 53 (39) | 43 (31) | 94 (69) | 65 (100%) | 62 (95%) | 3 (4·6%) |
| 2010 | 80 (100) | 34 (43) | 32 (40) | 48 (60) | 43 (100%) | 39 (91%) | 4 (9·3%) |
| 2011 | 44 (100) | 31 (70) | 31 (70) | 13 (30) | 15 (100%) | 13 (87%) | 2 (13%) |
| 2012 | 75 (100) | 48 (64) | 48 (64) | 27 (36) | 49 (100%) | 48 (98%) | 1 (2·0%) |
| 2013 | 45 (100) | 19 (42) | 19 (42) | 26 (58) | 20 (100%) | 18 (90%) | 2 (10%) |
| 2014 | 18 (100) | 17 (94) | 5 (28) | 13 (72) | 7 (100%) | 1 (14%) | 6 (86%) |
| 2015 | 68 (100) | 30 (44) | 30 (44) | 38 (56) | 12 (100%) | 8 (67%) | 4 (33%) |
| 2016 | 2 (100) | 2 (100) | 1 (50) | 1 (50) | 7 (100%) | 3 (43%) | 4 (57%) |
| 2017 | 14 (100) | 12 (86) | 4 (29) | 10 (71) | 0 (0%) | 0 (0%) | 0 (0%) |
| 2018 | 3 (100) | 3 (100) | 0 (0) | 3 (100) | 2 (100%) | 1 (50%) | 1 (50%) |
| 2019 | 203 (100) | 69 (34) | 68 (33) | 135 (67) | 84 (100%) | 47 (56%) | 37 (44%) |
| 2020 | 12 (100) | 11 (92) | 8 (67) | 4 (33) | 3 (100%) | 1 (33%) | 2 (67%) |
| 2021 | 20 (100) | 19 (95) | 7 (35) | 13 (65) | 0 (0%) | 0 (0%) | 0 (0%) |

[1]Typed: DENV serotype determined by RT-PCR (symptomatic infection) or EDIII-MMBA (inapparent infection).
[2]Missing serotype information. Stochastic imputation was applied to deal with missingness.

**Table S3. Imputation model performance on inapparent infections based on the experimentally serotyped data**

| Serotype | Prevalence | Accuracy | Sensitivity | Specificity | Overall accuracy | Overall kappa |
|---|---|---|---|---|---|---|
| DENV1 | 0·29 | 0·79 | 0·65 | 0·93 | 0·78 (0·74-0·82) | 0·66 |
| DENV2 | 0·47 | 0·84 | 0·85 | 0·83 | | |
| DENV3 | 0·22 | 0·89 | 0·86 | 0·91 | | |
| DENV4 | 0·02 | 0·66 | 0·33 | 0·99 | | |

**Table S4. Imputation model performance on symptomatic infections based on the experimentally serotyped data**

| Serotype | Prevalence | Accuracy | Sensitivity | Specificity | Overall accuracy | Overall kappa |
|---|---|---|---|---|---|---|
| DENV1 | 0·28 | 0·87 | 0·77 | 0·97 | 0·89 (0·85-0·92) | 0·83 |
| DENV2 | 0·35 | 0·94 | 0·92 | 0·96 | | |
| DENV3 | 0·37 | 0·93 | 0·96 | 0·90 | | |

**Table S5. Actual vs predicted inapparent DENV infections based only on the experimentally serotyped data**

| | | DENV1 | | DENV2 | | DENV3 | | DENV4 | |
|---|---|---|---|---|---|---|---|---|---|
| Year | n | Actual | Predicted | Actual | Predicted | Actual | Predicted | Actual | Predicted |
| 2004 | 40 | 17 | 19 | 17 | 18 | 1 | 0 | 4 | 4 |
| 2005 | 39 | 14 | 1 | 37 | 22 | 2 | 1 | 1 | 0 |
| 2006 | 13 | 3 | 0 | 13 | 10 | 0 | 0 | 0 | 0 |
| 2007 | 23 | 5 | 1 | 22 | 16 | 2 | 0 | 0 | 0 |
| 2008 | 14 | 4 | 7 | 2 | 4 | 5 | 5 | 1 | 0 |
| 2009 | 43 | 10 | 0 | 1 | 8 | 25 | 42 | 0 | 0 |
| 2010 | 32 | 0 | 0 | 2 | 6 | 26 | 30 | 0 | 0 |
| 2011 | 31 | 3 | 0 | 3 | 4 | 23 | 28 | 1 | 0 |
| 2012 | 48 | 45 | 48 | 0 | 1 | 2 | 0 | 0 | 0 |
| 2013 | 19 | 15 | 19 | 0 | 3 | 1 | 0 | 0 | 0 |
| 2014 | 5 | 1 | 0 | 1 | 1 | 2 | 2 | 1 | 2 |
| 2015 | 30 | 0 | 0 | 30 | 27 | 2 | 0 | 1 | 0 |
| 2016 | 1 | 0 | 0 | 1 | 1 | 0 | 0 | 0 | 0 |
| 2017 | 4 | 2 | 3 | 1 | 1 | 1 | 0 | 0 | 0 |
| 2019 | 68 | 0 | 0 | 68 | 68 | 0 | 0 | 0 | 0 |
| 2020 | 8 | 1 | 0 | 8 | 7 | 0 | 0 | 0 | 0 |
| 2021 | 7 | 4 | 5 | 2 | 3 | 0 | 0 | 0 | 0 |

**Table S6. Actual vs predicted symptomatic DENV infections based only on the experimentally serotyped data**

| | | DENV1 | | DENV2 | | DENV3 | |
|---|---|---|---|---|---|---|---|
| Year | n | Actual | Predicted | Actual | Predicted | Actual | Predicted |
| 2004 | 6 | 6 | 3 | 3 | 0 | 0 | 0 |
| 2005 | 20 | 10 | 8 | 12 | 10 | 0 | 0 |
| 2006 | 5 | 1 | 0 | 5 | 4 | 0 | 0 |
| 2007 | 23 | 1 | 0 | 23 | 22 | 0 | 0 |
| 2008 | 16 | 0 | 0 | 1 | 2 | 14 | 15 |
| 2009 | 62 | 8 | 3 | 0 | 1 | 53 | 59 |
| 2010 | 39 | 1 | 0 | 3 | 6 | 32 | 36 |
| 2011 | 13 | 3 | 0 | 0 | 0 | 10 | 13 |
| 2012 | 48 | 46 | 48 | 0 | 1 | 1 | 0 |
| 2013 | 18 | 11 | 12 | 0 | 2 | 5 | 6 |
| 2014 | 1 | 0 | 0 | 0 | 0 | 1 | 1 |
| 2015 | 8 | 0 | 0 | 8 | 8 | 0 | 0 |
| 2016 | 3 | 0 | 0 | 3 | 3 | 0 | 0 |
| 2018 | 1 | 0 | 0 | 1 | 1 | 0 | 0 |
| 2019 | 47 | 0 | 0 | 47 | 47 | 0 | 0 |
| 2020 | 1 | 0 | 0 | 1 | 1 | 0 | 0 |

### Table S7. Significance of fisher tests

| Cohort year | Imputed dataset | | Typed dataset | |
|---|---|---|---|---|
| | p.value[1] | p.star[2] | p.value | p.star |
| 2004 | 0·0105 | * | 0·0665 | ns |
| 2005 | 0·7686 | ns | 0·6877 | ns |
| 2006 | 1·0000 | ns | 1·0000 | ns |
| 2007 | 0·0205 | * | 0·0535 | · |
| 2008 | 0·0025 | ** | 0·0065 | ** |
| 2009 | 0·0005 | *** | 0·0010 | *** |
| 2010 | 0·2229 | ns | 0·8646 | ns |
| 2011 | 0·1959 | ns | 0·4423 | ns |
| 2012 | 1·0000 | ns | 1·0000 | ns |
| 2013 | 0·1894 | ns | 0·2179 | ns |
| 2014 | 0·1299 | ns | 1·0000 | ns |
| 2015 | 0·2639 | ns | 1·0000 | ns |
| 2016 | 1·0000 | ns | - | - |
| 2018 | 1·0000 | ns | - | - |
| 2019 | 0·0755 | ns | - | - |
| 2020 | 0·6082 | ns | 1·0000 | ns |

[1] p-value as determined by fisher test.
[2] *, p<0·05; **, p<0·01; ***, p<0·005.

### Table S8. Overall incidence of primary DENV infections by serotype.

| Serotype | Incidence* | Upper 95% CI | Lower 95% CI |
|---|---|---|---|
| DENV1 | 2·70 | 3·03 | 2·40 |
| DENV2 | 3·71 | 4·01 | 3·42 |
| DENV3 | 2·66 | 2·98 | 2·37 |

**This analysis excludes years when no symptomatic infections were reported. This is the result of intercept-only models. Incidence was calculated as the number of cases per serotype over the total population at risk (DENV-naïve individuals) times 100.*

### Table S9. Risk of dengue infections and percent symptomatic given primary infection by dengue serotype over time, based on model predictions across imputations excluding years with low symptomatic circulation (n < 10).

| | DENV1 | | DENV2 | | DENV3 | |
|---|---|---|---|---|---|---|
| Year | Incidence[1] | Percent symptomatic | Incidence[1] | Percent symptomatic | Incidence[1] | Percent symptomatic |
| 2004 | 4 (3-5) | 15 (8-28) | - | - | - | - |
| 2005 | 4 (3-5) | 19 (10-32) | 6 (5-7) | 12 (7-22) | - | - |
| 2006 | 1 (1-2) | 6 (1-35) | 4 (3-5) | 6 (2-16) | - | - |
| 2007 | 1 (1-2) | 4 (1-26) | 5 (4-7) | 26 (18-36) | - | - |
| 2008 | - | - | 1 (1-2) | 8 (2-29) | 2 (2-3) | 38 (23-55) |
| 2009 | 2 (1-3) | 22 (11-39) | 2 (1-3) | 3 (0-20) | 7 (6-9) | 42 (34-51) |
| 2010 | - | - | 2 (1-3) | 25 (11-46) | 6 (5-7) | 37 (28-48) |
| 2011 | - | - | - | - | 3 (2-4) | 25 (15-40) |
| 2012 | 6 (5-7) | 40 (32-49) | - | - | - | - |
| 2013 | 2 (2-3) | 26 (16-41) | - | - | - | - |
| 2014 | - | - | - | - | 1 (0-1) | 44 (22-69) |
| 2015 | - | - | 3 (2-4) | 16 (9-27) | - | - |
| 2016 | - | - | - | - | - | - |
| 2017 | - | - | - | - | - | - |
| 2018 | - | - | - | - | - | - |
| 2019 | - | - | 17 (15-19) | 29 (24-35) | - | - |
| 2020 | - | - | 1 (0-1) | 24 (8-54) | - | - |
| 2021 | - | - | - | - | - | - |

[1]Incidence per 100 naive individuals

**Table S10. Odds ratios of symptomatic primary infection given DENV1 infection over time**

| Year | Pooled OR | Std error | Low 95% CI | High 95% CI | P-value |
|---|---|---|---|---|---|
| 2004 | 0.26 | 0.44 | 0.11 | 0.62 | 0.002 |
| 2005 | 0.34 | 0.41 | 0.15 | 0.76 | 0.009 |
| 2006 | 0.1 | 1.07 | 0.01 | 0.84 | 0.034 |
| 2007 | 0.07 | 1.05 | 0.01 | 0.54 | 0.011 |
| 2009 | 0.42 | 0.46 | 0.17 | 1.04 | 0.061 |
| 2012 | ref | ref | ref | ref | ref |
| 2013 | 0.54 | 0.39 | 0.25 | 1.16 | 0.11 |

**Table S11. Odds ratios of symptomatic primary infection given DENV2 infection over time**

| Year | Pooled | Std error | Low 95% CI | High 95% CI | P-value |
|---|---|---|---|---|---|
| 2005 | 0.34 | 0.36 | 0.17 | 0.7 | 0.003 |
| 2006 | 0.16 | 0.54 | 0.06 | 0.47 | < 0.001 |
| 2007 | 0.84 | 0.28 | 0.48 | 1.45 | 0.53 |
| 2008 | 0.22 | 0.76 | 0.05 | 0.99 | 0.049 |
| 2009 | 0.08 | 01.03 | 0.01 | 0.63 | 0.016 |
| 2010 | 0.79 | 0.51 | 0.29 | 2.14 | 0.65 |
| 2015 | 0.47 | 0.34 | 0.24 | 0.92 | 0.028 |
| 2019 | ref | ref | ref | ref | ref |
| 2020 | 0.76 | 0.68 | 0.2 | 2.89 | 0.68 |

**Table S12. Odds ratios of symptomatic primary infection given DENV3 infection over time**

| Year | Pooled | Std error | Low 95% CI | High 95% CI | P-value |
|---|---|---|---|---|---|
| 2008 | 0.84 | 0.4 | 0.38 | 1.86 | 0.67 |
| 2009 | ref | ref | ref | ref | ref |
| 2010 | 0.81 | 0.29 | 0.46 | 1.42 | 0.46 |
| 2011 | 0.47 | 0.39 | 0.21 | 01.01 | 0.054 |
| 2014 | 01.08 | 0.57 | 0.35 | 3.31 | 0.89 |

### Table S13. Sensitivity analyses

| Sensitivity analysis | Serotype | Outcome | Event | Infected | Percent | Pooled OR (95% CI) | P-value |
|---|---|---|---|---|---|---|---|
| No imputation - experimentally typed data | DENV1 | Symptomatic given infection | 87 | 211 | 41·23 | Ref. | Ref. |
| | DENV2 | | 108 | 308 | 35·06 | 0·78 (0·44 to 1·39) | 0·40 |
| | DENV3 | | 116 | 208 | 55·77 | 1·83 (1·01 to 3·34) | 0·046 |
| | DENV1 | Severe given infection | 10 | 211 | 4·74 | Ref | Ref |
| | DENV2 | | 9 | 308 | 2·92 | 1·12 (0·24 to 4·93) | 0·89 |
| | DENV3 | | 26 | 208 | 12·5 | 5·61 (1·69 to 21·17) | 0·0072 |
| Polytomous regression framework (imputing DENV-infected with no EDIII) | DENV1 | Symptomatic given infection | 94 | 337 | 27·89 | Ref | Ref |
| | DENV2 | | 157 | 663 | 23·68 | 0·83 (0·50 to 1·36) | 0·455 |
| | DENV3 | | 131 | 318 | 41·19 | 1·81 (1·08 to 3·05) | 0·025 |
| | DENV1 | Severe given infection | 10 | 337 | 2·97 | Ref | Ref |
| | DENV2 | | 11 | 663 | 1·66 | 1·19 (0·29 to 4·90) | 0·808 |
| | DENV3 | | 27 | 318 | 8·49 | 5·60 (1·69 to 18·52) | 0·005 |
| Classification trees imputation framework | DENV1 | Symptomatic given infection | 96 | 418 | 22·97 | Ref | Ref |
| | DENV2 | | 159 | 781 | 20·36 | 0·93 (0·57 to 1·54) | 0·787 |
| | DENV3 | | 126 | 366 | 34·43 | 1·97 (1·18 to 3·28) | 0·01 |
| | DENV1 | Severe given infection | 10 | 418 | 2·39 | Ref | Ref |
| | DENV2 | | 11 | 781 | 1·41 | 1·38 (0·34 to 5·56) | 0·651 |
| | DENV3 | | 27 | 366 | 7·38 | 6·39 (1·96 to 20·88) | 0·002 |
| Random forest framework | DENV1 | Symptomatic given infection | 96 | 426 | 22·54 | Ref | Ref |
| | DENV2 | | 155 | 768 | 20·18 | 0·91 (0·56 to 1·46) | 0·682 |
| | DENV3 | | 130 | 368 | 35·33 | 2·18 (1·32 to 3·59) | 0·002 |
| | DENV1 | Severe given infection | 10 | 426 | 2·35 | Ref | Ref |
| | DENV2 | | 11 | 768 | 1·43 | 1·31 (0·35 to 4·84) | 0·687 |
| | DENV3 | | 27 | 368 | 7·34 | 5·94 (1·91 to 18·50) | 0·002 |
| Predictive mean matching imputation framework | DENV1 | Symptomatic given infection | 97 | 413 | 23·49 | Ref | Ref |
| | DENV2 | | 151 | 774 | 19·51 | 0·91 (0·54 to 1·55) | 0·729 |
| | DENV3 | | 135 | 377 | 35·81 | 2·14 (1·10 to 4·17) | 0·025 |
| | DENV1 | Severe given infection | 10 | 413 | 2·42 | Ref | Ref |
| | DENV2 | | 11 | 774 | 1·42 | 1·37 (0·38 to 4·89) | 0·63 |
| | DENV3 | | 27 | 377 | 7·16 | 6·35 (1·86 to 21·66) | 0·003 |
| Polytomous regression (outcome models adjusted by age and sex) | DENV1 | Symptomatic given infection | 94 | 402 | 23·38 | Ref | Ref |
| | DENV2 | | 157 | 782 | 20·08 | 0·78 (0·46 to 1·32) | 0·357 |
| | DENV3 | | 131 | 363 | 36·09 | 2·36 (1·37 to 4·09) | 0·002 |
| | DENV1 | Severe given infection | 10 | 402 | 2·49 | Ref | Ref |
| | DENV2 | | 11 | 782 | 1·41 | 1·20 (0·23 to 6·37) | 0·83 |
| | DENV3 | | 27 | 363 | 7·44 | 10·02 (2·19 to 45·83) | 0·003 |
| Polytomous regression framework (serotype ~ year) | DENV1 | Symptomatic given infection | 93 | 418 | 22·25 | Ref | Ref |
| | DENV2 | | 158 | 761 | 20·76 | 0·98 (0·60 to 1·60) | 0·933 |
| | DENV3 | | 131 | 377 | 34·75 | 2·09 (1·25 to 3·50) | 0·005 |
| | DENV1 | Severe given infection | 10 | 418 | 2·39 | Ref | Ref |
| | DENV2 | | 11 | 761 | 1·45 | 1·39 (0·35 to 5·59) | 0·639 |
| | DENV3 | | 27 | 377 | 7·16 | 6·30 (1·93 to 20·56) | 0·002 |

| Sensitivity analysis | Serotype | Outcome | Event | Infected | Percent | Pooled OR (95% CI) | P-value |
|---|---|---|---|---|---|---|---|
| Polytomous regression framework (serotype ~ region*year) | DENV1 | Symptomatic given infection | 93 | 415 | 22·41 | Ref | Ref |
| | DENV2 | | 158 | 764 | 20·68 | 0·90 (0·55 to 1·48) | 0·688 |
| | DENV3 | | 131 | 375 | 34·93 | 2·10 (1·25 to 3·51) | 0·005 |
| | DENV1 | Severe given infection | 10 | 415 | 2·41 | Ref | Ref |
| | DENV2 | | 11 | 764 | 1·44 | 1·40 (0·35 to 5·59) | 0·634 |
| | DENV3 | | 27 | 375 | 7·2 | 6·28 (1·93 to 20·41) | 0·002 |
| Polytomous regression framework (serotype ~ region*year + age) | DENV1 | Symptomatic given infection | 92 | 403 | 22·83 | Ref | Ref |
| | DENV2 | | 154 | 722 | 21·33 | 1·01 (0·62 to 1·66) | 0·955 |
| | DENV3 | | 134 | 368 | 36·41 | 2·40 (1·45 to 3·97) | <0·001 |
| | DENV1 | Severe given infection | 10 | 403 | 2·48 | Ref | Ref |
| | DENV2 | | 11 | 722 | 1·52 | 1·43 (0·37 to 5·47) | 0·603 |
| | DENV3 | | 27 | 368 | 7·34 | 6·34 (2·06 to 19·55) | 0·001 |
| Ignoring DENV4 infections | DENV1 | Symptomatic given infected | 94 | 419 | 22·43 | Ref | Ref |
| | DENV2 | | 157 | 797 | 19·7 | 0·87 (0·54 to 1·41) | 0·58 |
| | DENV3 | | 131 | 375 | 34·93 | 2·09 (1·27 to 3·43) | 0·004 |
| | DENV1 | Severe given infected | 10 | 419 | 2·39 | Ref | Ref |
| | DENV2 | | 11 | 797 | 1·38 | 1·22 (0·31 to 4·79) | 0·775 |
| | DENV3 | | 27 | 375 | 7.2 | 6·45 (1·97 to 21·15) | 0·002 |
| Constraining study period to 2004-2016 (Prior to ZIKV emergence) | DENV1 | Symptomatic given infected | 93 | 379 | 24·54 | Ref | Ref |
| | DENV2 | | 63 | 461 | 13·67 | 0·77 (0·47 to 1·27) | 0·312 |
| | DENV3 | | 131 | 360 | 36·39 | 2·12 (1·26 to 3·56) | 0·005 |
| | DENV1 | Severe given infected | 10 | 379 | 2·64 | Ref | Ref |
| | DENV2 | | 5 | 461 | 1·08 | 0·99 (0·24 to 4·14) | 0·987 |
| | DENV3 | | 27 | 360 | 7·50 | 6·41 (1·93 to 21·35) | 0·002 |
| Removing the 76 samples from the EDIII-MMBA evaluation set | DENV1 | Symptomatic given infected | 94 | 374 | 25·13 | Ref | Ref |
| | DENV2 | | 157 | 746 | 21·05 | 0·83 (0·50 to 1·38) | 0·482 |
| | DENV3 | | 131 | 346 | 37·86 | 2·12 (1·25 to 3·59) | 0·006 |
| | DENV1 | Severe given infected | 10 | 374 | 2·67 | Ref | Ref |
| | DENV2 | | 11 | 746 | 1·47 | 1·16 (0·29 to 4·73) | 0·833 |
| | DENV3 | | 27 | 346 | 7·80 | 6·33 (1·83 to 21·83) | 0·004 |

### Table S14. Outcome of primary infections

| Serotype | Inapparent | Symptomatic | | | |
|---|---|---|---|---|---|
| | | Overall | DwoWS | DwWS | SD |
| **DENV1** | 76.8% (72.1-80.8%) | 23.2% (19.2-27.8%) | 11.2% (8.4-14.8%) | 9.5% (7.0-12.9%) | 2.5% (1.3-4.6%) |
| **DENV2** | 79.9% (76.8-82.6%) | 20.1% (17.4-23.2%) | 14.1% (11.8-16.8%) | 4.6% (3.3-6.3%) | 1.4% (0.8-2.6%) |
| **DENV3** | 63.9% (58.6-68.9%) | 36.1% (31.1-41.4%) | 16.4% (12.8-20.7%) | 12.3% (9.3-16.2%) | 7.4% (5.1-10.6%) |

### Table S15. Genotype of Nicaraguan DENV serotypes.

| Serotype | Genotype |
|---|---|
| DENV1 | Genotype V (American/East African and Asian) |
| DENV2 | Genotype IIIb (Southeast Asian-American) |
| DENV3 | Genotype III (Indian-subcontinent) |

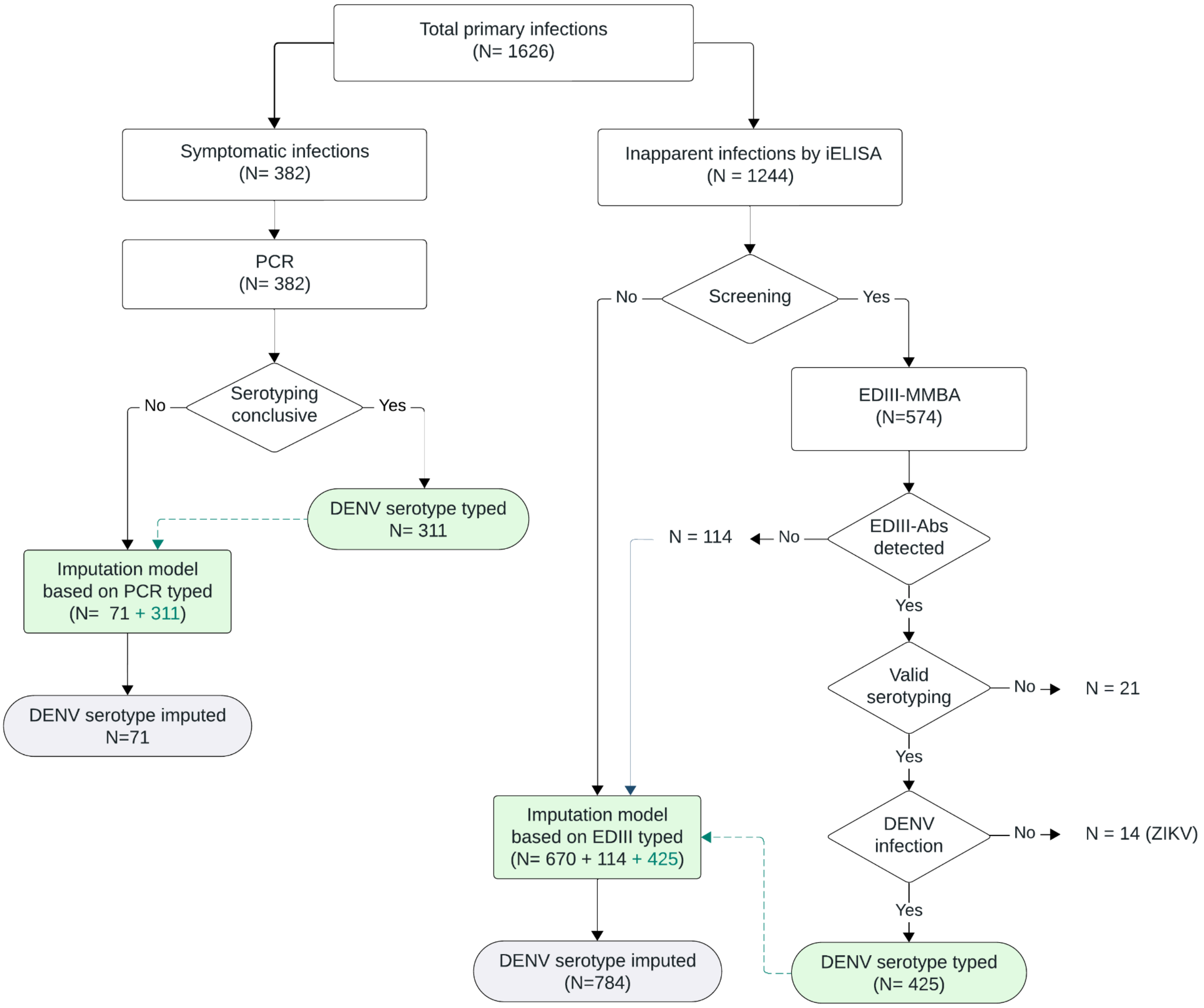


**Figure S1. Flowchart of the screening and serotyping process to study the epidemiology of primary DENV infections in the PDCS**

Serotyping inconclusive refers to participants with primary symptomatic infection via MAC-ELISA and/or iELISA, with no serotyping data by RT-PCR due to negative, unclear results or low viremia. Valid serotyping refers to participants for whom conclusive serotyping was performed via EDIII-MMBA according to the rules detailed in the Supplementary Appendix, section “Typing of primary inapparent infection based on EDIII-MMBA”.

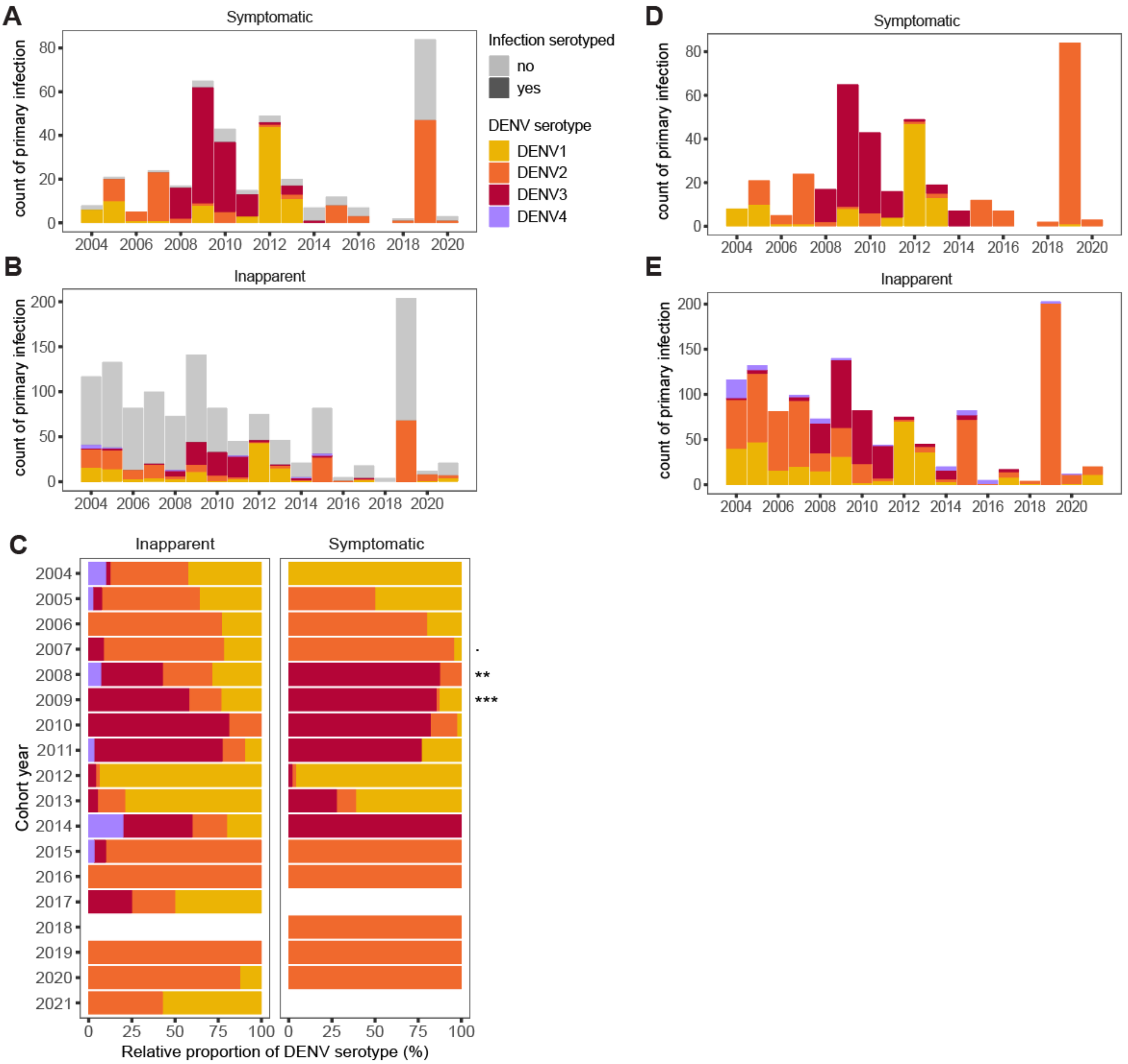


**Figure S2. Distribution of serotyped, non-serotyped, and imputed symptomatic and inapparent primary DENV from 2004 to 2021 in the PDCS**

**(A)** Yearly counts of symptomatic primary DENV infections by DENV serotype as serotyped by rRT-PCR. **(B)** Yearly counts of inapparent primary DENV infections by DENV serotype as serotyped by EDIII-MMBA. **(C)** Relative distribution of DENV serotypes circulating yearly in inapparent and symptomatic primary DENV infections as serotyped by rRT-PCR and EDIII-MMBA. **(D)** Pooled yearly counts of symptomatic primary DENV infections by DENV serotype across multiple imputations **(E)** Pooled yearly counts of inapparent primary DENV infections by DENV serotype across multiple imputations. p-value, p=0·05; **, p<0·01; ***, p<0·005.

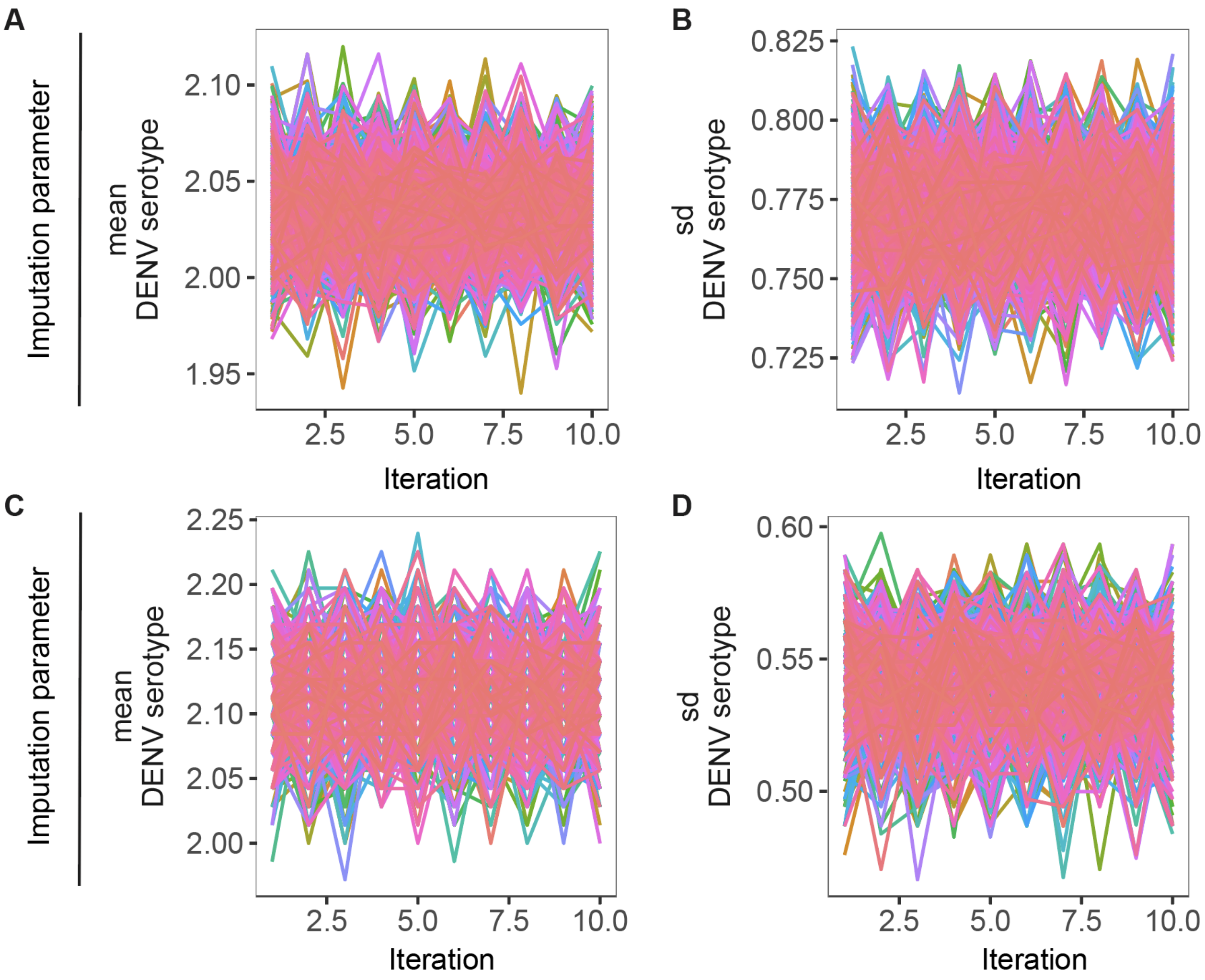


**Figure S3. Trace plots for the final imputation models**

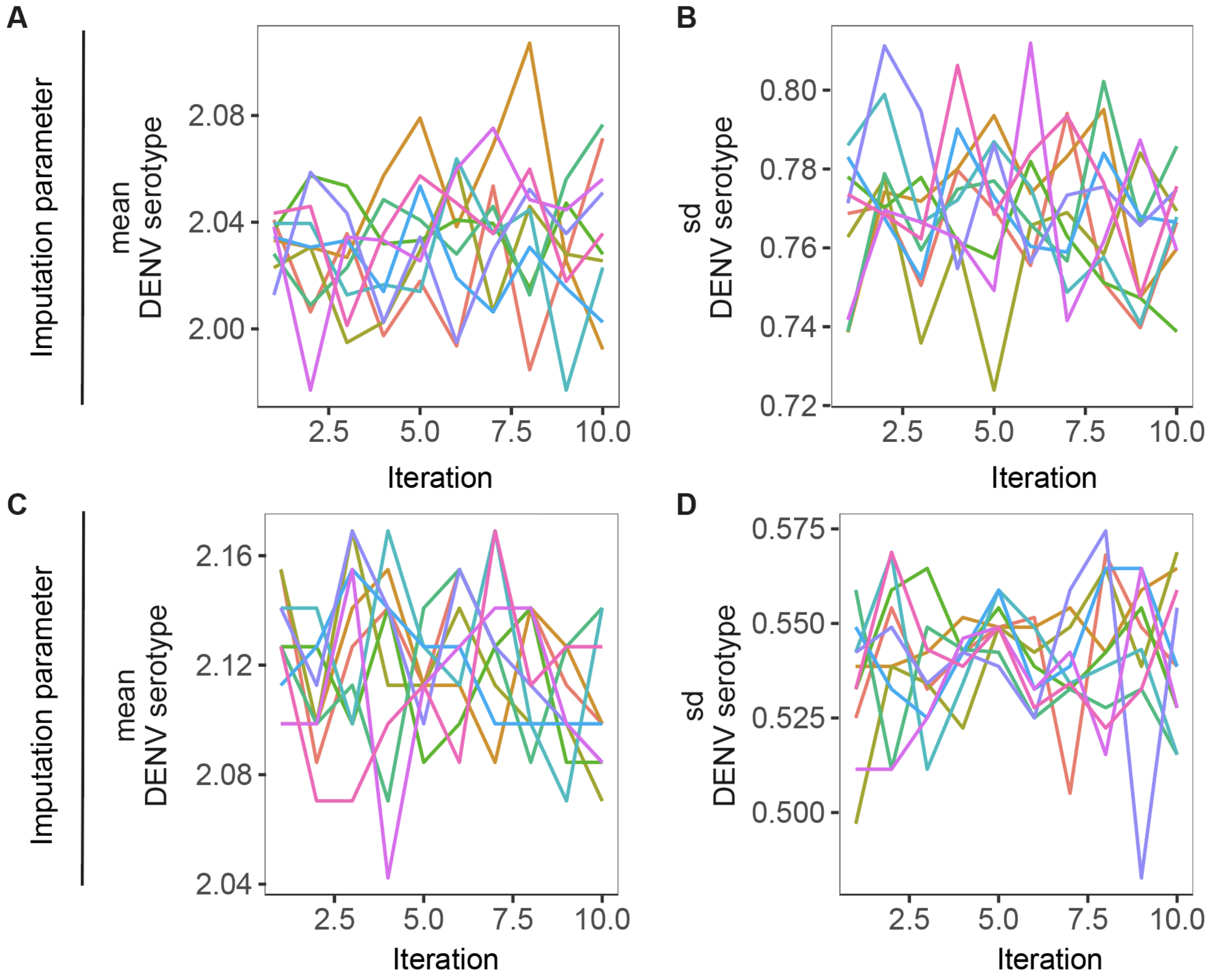


**Figure S4. Trace plots for the final imputation framework based on only 10 simulations**

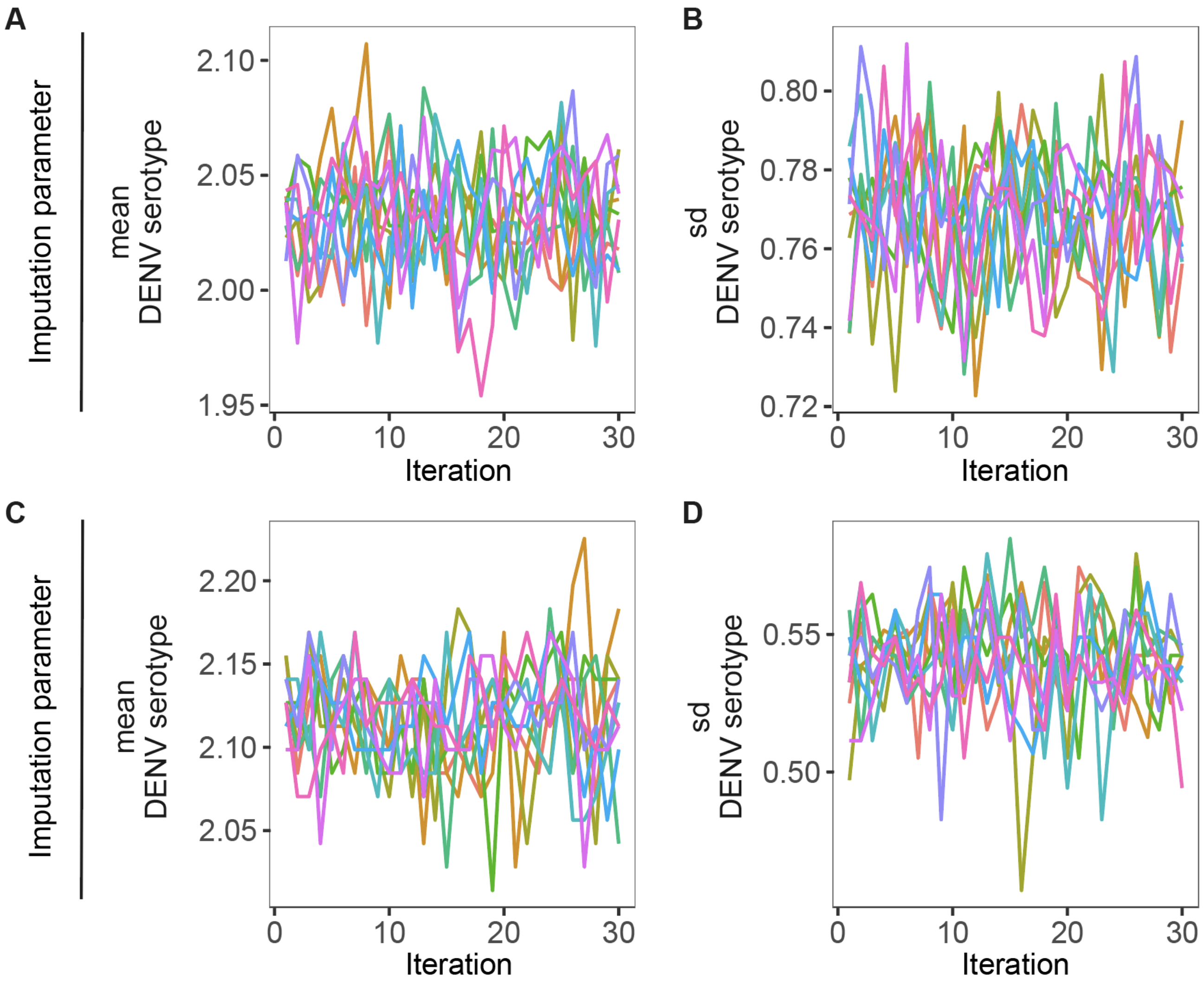


**Figure S5. Trace plots for the final imputation framework based on only 10 simulations and 30 iterations**

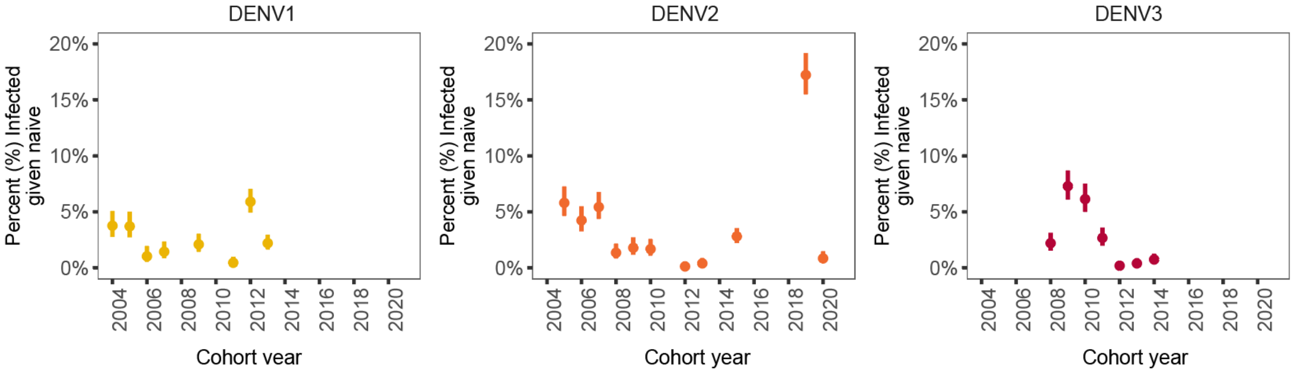


**Figure S6. Temporal dynamics of overall primary DENV infections by serotype from 2004 to 2021 in the PDCS**

Predictions and 95% CI of percent of primary DENV infections by infection outcome and serotype among the naive population, as indicated in left, middle, and right panels across imputations. Years without symptomatic circulation were not included.

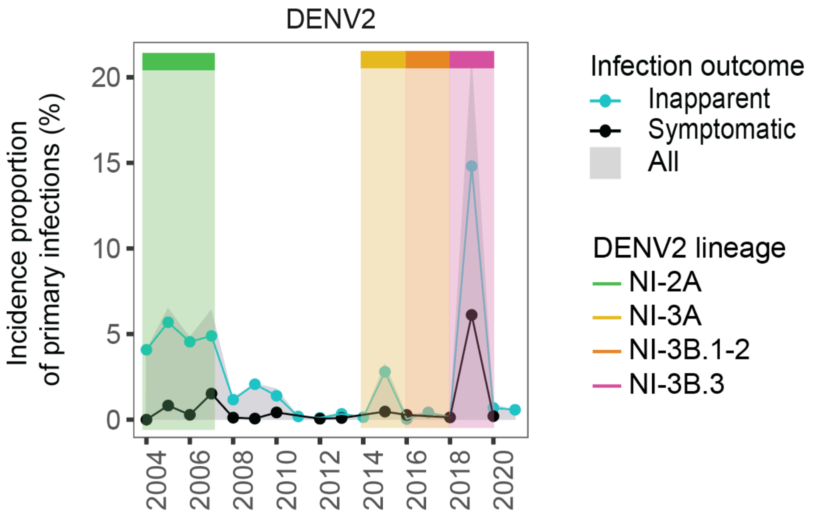


**Figure S7. Temporal shift in Nicaraguan DENV2 lineages**

Annual incidence proportion of primary DENV2 infections alongside the temporal evolution of DENV2 lineages in Nicaragua, mapped onto the respective years. Detailed descriptions of the Nicaraguan DENV2 lineages and pertinent information regarding amino acid mutations can be found in Thongsripong et al[17].

### Supplementary References


1 Kuan G, Gordon A, Avilés W, *et al.* The Nicaraguan pediatric dengue cohort study: study design, methods, use of information technology, and extension to other infectious diseases. *Am J Epidemiol* 2009; **170**: 120–9.

2 Gordon A, Kuan G, Aviles W, *et al.* The Nicaraguan pediatric influenza cohort study: design, methods, use of technology, and compliance. *BMC Infect Dis* 2015; **15**: 504.

3 Zambrana JV, Bustos Carrillo F, Burger-Calderon R, *et al.* Seroprevalence, risk factor, and spatial analyses of Zika virus infection after the 2016 epidemic in Managua, Nicaragua. *Proc Natl Acad Sci U S A* 2018; **115**: 9294–9.

4 Gordon A, Kuan G, Mercado JC, *et al.* The Nicaraguan pediatric dengue cohort study: incidence of inapparent and symptomatic dengue virus infections, 2004-2010. *PLoS Negl Trop Dis* 2013; **7**: e2462.

5 Waggoner JJ, Gresh L, Mohamed-Hadley A, *et al.* Single-Reaction Multiplex Reverse Transcription PCR for Detection of Zika, Chikungunya, and Dengue Viruses. *Emerg Infect Dis* 2016; **22**: 1295–7.

6 Lanciotti RS, Calisher CH, Gubler DJ, Chang GJ, Vorndam AV. Rapid detection and typing of dengue viruses from clinical samples by using reverse transcriptase-polymerase chain reaction. *J Clin Microbiol* 1992; **30**: 545–51.

7 Balmaseda A, Sandoval E, Pérez L, Gutiérrez CM, Harris E. Application of molecular typing techniques in the 1998 dengue epidemic in Nicaragua. *Am J Trop Med Hyg* 1999; **61**: 893–7.

8 Waggoner JJ, Abeynayake J, Sahoo MK, *et al.* Single-reaction, multiplex, real-time rt-PCR for the detection, quantitation, and serotyping of dengue viruses. *PLoS Negl Trop Dis* 2013; **7**: e2116.

9 Balmaseda A, Zambrana JV, Collado D, *et al.* Comparison of Four Serological Methods and Two Reverse Transcription-PCR Assays for Diagnosis and Surveillance of Zika Virus Infection. *J Clin Microbiol* 2018; **56**. DOI:10.1128/JCM.01785-17.

10 Balmaseda A, Guzmán MG, Hammond S, *et al.* Diagnosis of dengue virus infection by detection of specific immunoglobulin M (IgM) and IgA antibodies in serum and saliva. *Clin Diagn Lab Immunol* 2003; **10**: 317–22.

11 Gordon A, Gresh L, Ojeda S, *et al.* Prior dengue virus infection and risk of Zika: A pediatric cohort in Nicaragua. *PLoS Med* 2019; **16**: e1002726.

12 Katzelnick LC, Gresh L, Halloran ME, *et al.* Antibody-dependent enhancement of severe dengue disease in humans. *Science* 2017; **358**: 929–32.

13 World Health Organization. Dengue guidelines for diagnosis, treatment, prevention and control : new edition. World Health Organization, 2009 https://iris.who.int/handle/10665/44188 (accessed March 2, 2024).

14 Bos S, Graber AL, Cardona-Ospina JA, *et al.* Protection against symptomatic dengue infection by neutralizing antibodies varies by infection history and infecting serotype. *Nat Commun* 2024; **15**: 382.

15 Fernández RJ, Vázquez S. Serological diagnosis of dengue by an ELISA inhibition method (EIM). *Mem Inst Oswaldo Cruz* 1990; **85**: 347–51.

16 Reed LJ, Muench H. A SIMPLE METHOD OF ESTIMATING FIFTY PER CENT ENDPOINTS. *Am J Epidemiol* 1938; **27**: 493–7.

17 Thongsripong P, Edgerton SV, Bos S, *et al.* Phylodynamics of dengue virus 2 in Nicaragua leading

up to the 2019 epidemic reveals a role for lineage turnover. *BMC Ecol Evol* 2023; **23**: 58.
